# Maternal cardiovascular and haematological complications alter the risk associations between environmental exposure and adverse pregnancy outcomes

**DOI:** 10.1101/2023.11.15.23298338

**Authors:** Jason Sun, Haiyang Tang, Huan Zhao, Qingyi Xiang, Yijia Tian, Kim Robin van Daalen, Kun Tang, Evelyn Xiu-Ling Loo, Lynette P Shek, Alexander T Archibald, Wei Xu, Yuming Guo, Xiaoxia Bai, the Zhejiang Environmental and Birth Health Research Alliance (ZEBRA) collaborative group

## Abstract

Given China’s recent introduction of the “three-child policy” in response to population ageing^1^, safeguarding perinatal health has become an urgent priority^2^. Previous epidemiological research seldom explored the risk factors of maternal cardiovascular and haematological diseases, or its impact on adverse pregnancy outcomes (APO). To fill the literature gap, here we conducted systematic epidemiological analyses on 121,090 pregnant women and their neonates from the ZEBRA Chinese prospective maternity cohort. We find that incremental exposure in PM_2.5_, O_3_, and green space modify the risks of APO, including congenital heart disease, by 11.2%, 7.8%, and –5.5%, respectively. Maternal cardiovascular and haematological complications during pregnancy significantly aggravate the risk of APO by 66.2%, and also modify the environment-APO risk associations by amplifying the hazards of air pollution and weakening the protective effect of greenness accessibility. Our research findings support the Sustainable Development Goals (e.g. SDG3)^3,4^ by providing first-hand epidemiological evidence and clinical guidance for protecting maternal and neonatal health.

---

Studies have assessed the effects of environmental exposures, including ambient air pollution, heatwave, and green space, on the risk of cardiovascular morbidity and mortality^5-10^. However, research that focused on the pregnant woman and the neonate, two groups of vulnerable population, is still limited. Literature has also reported strong associations between maternal exposure to various environmental factors and multiple types of APO^11-16^, but other pregnancy abnormalities (e.g. neonatal congenital heart disease, CHD) other than preterm birth (PTB), term low birth weight (LBW), and stillbirth have seldom been analysed. It is noteworthy that women with pregnancy-induced cardiovascular diseases (CVDs) have an increased risk of developing hypertension in later years^17,18^. Research has shown that women with a history of pre-eclampsia have an approximately four-fold higher incidence of stroke in later years^17^, as pre-eclampsia can cause maternal vascular remodelling^19^. For the neonates diagnosed with CHD, childhood mortality rates are rather high^20^; even with medical treatment, adults and adolescents often have lower exercise capability^21^, and a portion of women are even advised against carrying to pregnancy to term in fear of cardiac complications and even sudden death of the child^22^. Although highly sensitive screening tools are now being widely used for timely prenatal detection of CHD^23^, identifying risk factors to minimise the occurrence risk of CHD is always the ultimate pursuit.

Therefore, population-based epidemiological research on cardiovascular-related outcomes during the perinatal period holds significant public health implications. By systematic analyses of the medical records of pregnant women enrolled in the ZEBRA Chinese maternity cohort^24^, our current study aims to explore: 1) the risk associations between environmental exposures and pregnancy-induced CVDs and APO; 2) the mediating role of maternal cardiovascular symptoms (both primary and pregnancy-induced) in the environment-APO risk association; and 3) the early-stage predictability of APO integrating maternal diagnosis of CVDs. Besides contributing to filling the literature gap, the risk prediction models also provide the basis for further clinical validation and application. Protecting the mother’s health is not only an important element of gender equality, but also a reflection of civilisation. Ensuring the health of newborns is crucial as it directly impacts the lifelong well-being and human potential of the next generation, serving as a vital factor in advancing society.

## RESULTS

The ZEBRA (Zhejiang Environmental and Birth Health Research Alliance) maternity cohort recruited 137,392 Chinese pregnant women nationwide during 2013–2022, among which 121,090 pregnant women were included for epidemiological analyses (Fig. 1). There were 25,544 (21.1%) pregnant women diagnosed with a variety of cardiovascular and haematological complications, including 814 (0.7%) cases of heart disease, 3,760 (3.1%) cases of vascular disease, and 21,935 (18.1%) cases of haematological disease. Pregnancy-induced cardiovascular complications were found in 1,414 (1.2%) cases (Extended Data Fig. 1). After excluding 802 stillbirths, there were 9,501 cases of APOs among the remaining 124,025 live births (3,619 pairs of twins and 59 sets of triplets), including 6,604 (5.3%) PTB, 4,849 (3.9%) term LBW, 840 (0.7%) CHD, and 3,334 (2.7%) blood disorders (Extended Data Fig. 2). Detailed cohort profile statistics are listed in Extended Data Table 1, and medical history in Extended Data Table 2.

**Fig. 1.**
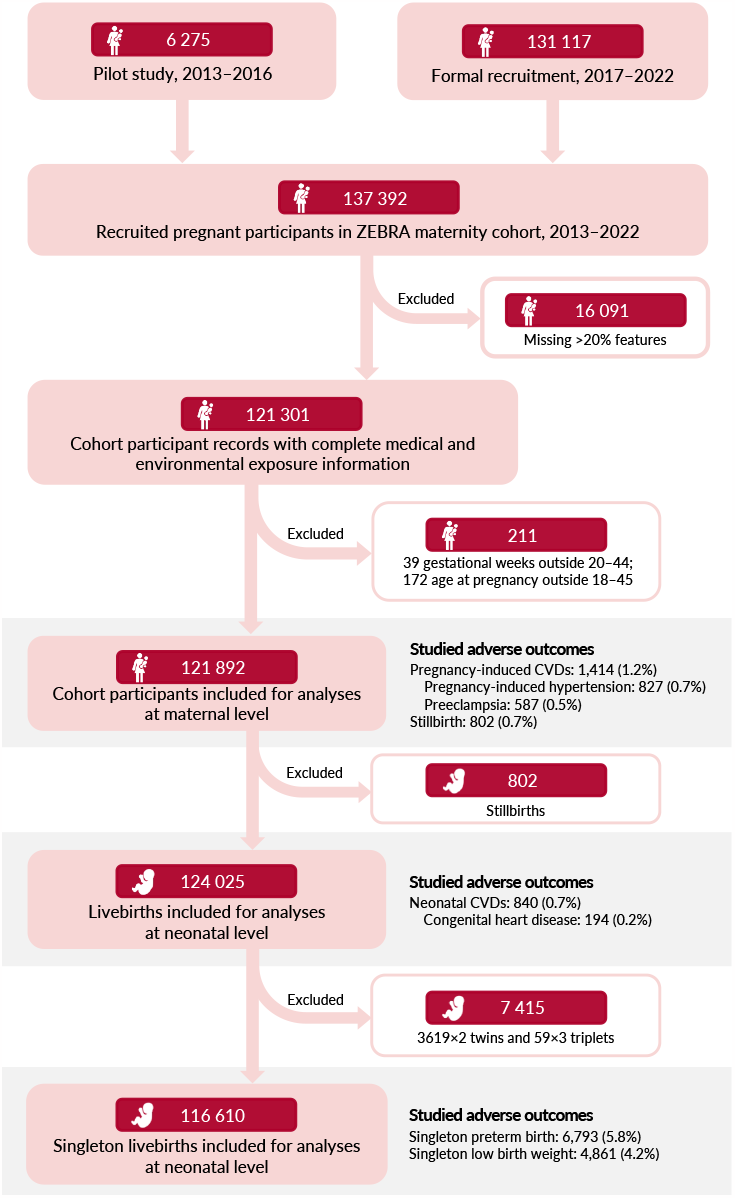
Recruitment flowchart of ZEBRA maternity cohort participants, 2013–2022. ZEBRA maternity cohort comprises a comprehensive dataset, including sociodemographic characteristics, medical history prior to conception, diagnoses during pregnancy, obstetric-related diagnoses during the perinatal period, physiological and biochemical parameters obtained through routine blood and urine tests, and a retrospective record of environmental exposure tracing. Participants who changed their place of residence during the 18-month period for analysis (from one year before conception to the end of the second trimester) or had records missing >20% of studied variables were excluded from the analysis. Additionally, participants with gestational durations <20 or >44 weeks and aged <18 or >45 years were censored to eliminate potential effects on pregnancy outcomes. Shaded participants represent the cohort members included in designed analyses investigating three epidemiological aspects: i) the influence of risk factors on the occurrence of maternal pregnancy-induced cardiovascular diseases and stillbirths; ii) the relationship between risk factors and neonatal cardiovascular disorders; and iii) the association between risk factors and obstetric anomalies in singleton neonates.

### Environmental risks on perinatal CVDs

To investigate the risk associations between environmental exposure and pregnancy-induced cardiovascular complications, we defined the eighteen month period starting from one year before conception till the end of the second trimester as the exposure window^25^. This yielded individual-level maternal exposure to ambient PM_2.5_, O_3_, and greenness as 40.3±7.4 μg/m^3^, 47.1±4.0 ppb, and 0.20±0.07 quantified in the enhanced vegetation index (EVI), respectively. Having adjusted for sociodemographic, physiological and behavioural characteristics, medical and disease history, and other environmental exposures (see details in Extended Data Table 3), the risk of pregnancy-induced cardiovascular diseases would increase by 7.3% (hazard ratio, HR=1.073, 95% CI: 1.064–1.082, *p*=7.83×10^−23^) with each 10-μg/m^3^ increase in PM_2.5_ exposure, increase by 2.7% (2.2–3.3%, *p*=1.78×10^−12^) with each 10-ppb incremental O_3_ exposure, and reduce by 3.6% (1.8–5.2%, *p*=1.47×10^−4^) with every 0.1-EVI additional green space exposure. Significant risk associations were observed for two specific subsets of cardiovascular diseases, that is, the pregnancy-induced hypertension and preeclampsia (Fig. 2).

**Fig. 2.**
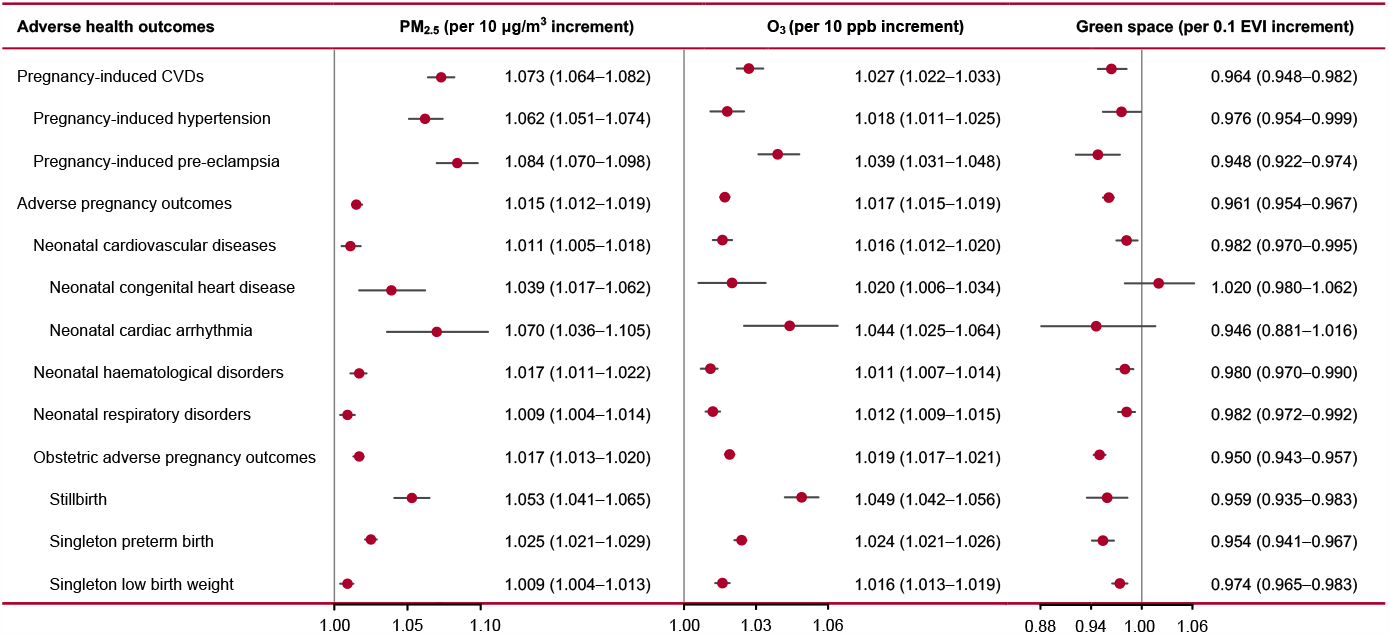
Risk association between environmental exposure and perinatal abnormalities. Epidemiological analyses encompass a variety of adverse health outcomes, including maternal pregnancy-induced cardiovascular diseases (CVDs) (excluding primary CVDs), neonatal cardiovascular and haematological disorders, as well as stillbirth, singleton preterm birth, and singleton low birth weight. Environmental exposure tracking covers PM_2.5_, O_3_, and greenness, to which hazard ratios (HR) with 95% confidence intervals (CI) were estimated sufficiently having adjusted for covariates for 10-μg/m^3^, 10-ppb, and 0.1-EVI incremental exposure, respectively. Subcategories of adverse health outcomes that did not exhibit significant risk associations with three environmental exposures were not included in the forest-plot. The indentation degree of listed subcategories of adverse health outcomes represents hierarchical tiers.

As for the neonatal cardiovascular diseases, it was found that for every 10-μg/m^3^ exposure increase in PM_2.5_, 10-ppb O_3_, and 0.1-EVI greenness, there would be 1.1% (0.5–1.8%, *p*=1.63×10^−3^), 1.6% (1.2–2.0%, *p*=1.79×10^−10^), and –1.8% (HR=0.982, 95% CI: 0.970–0.995, *p*=4.79×10^−3^) change in the incidence risks, respectively (Fig. 2). The neonatal cardiac anomalies had the most prominent associations with maternal PM_2.5_ and O_3_ exposures. The protective effect of maternal greenness exposure was not significant on the neonatal incidence of CHD Environmental exposures also demonstrated additional risks on other adverse perinatal outcomes such as stillbirth, PTB, term LBW, and respiratory disease, where the quantitative relationships showed monotonically increasing exposure-response tendencies (Extended Data Fig. 3).

Synergistic effect between PM_2.5_ and O_3_ exposure was observed as 4.2% (effect modification, EM=1.042, 95% CI: 1.022–1.062, *p*=3.88×10^−5^) for pregnancy-induced cardiovascular complications, indicating for each 10-ppb increase in maternal O_3_ exposure, the risk strength between PM_2.5_ exposure (by 10-μg/m^3^ increment) and cardiovascular incidence would rise by 4.2%, and *vice versa*. The synergistic hazard was more significant in the incidence risk of APOs (EM=1.054, 95% CI: 1.046–1.061, *p*=2.33×10^−23^). On the contrary, green space accessibility exhibited antagonistic effects with the two major ambient air pollutants, as –9.7% (EM=0.903, 95% CI: 0.883–0.923, *p*=2.41×10^−13^) on the risk association between PM_2.5_ and pregnancy-induced cardiovascular diseases, implying that with each 0.1-EVI incremental greenness exposure, the PM_2.5_-cardiovascular risk strength scaled in 10-μg/m^3^ increase of exposure would be compromised by 9.7%. The antagonism of greenness exposure on O_3_ was relatively weaker at –4.4% (EM=0.956, 95% CI: 0.938–0.973, *p*=2.56×10^−6^). The protective effect of greenness against the occurrence risk of APO was 8.6% (7.7–9.4%, *p*=2.08×10^−5^) and 4.7% (4.1–5.4%, *p*=2.15×10^−20^) for PM_2.5_ and O_3_ exposure, respectively. The foregoing analyses revealed unneglectable risks associated with ambient air pollution on the well-being of expectant mothers and neonates, particularly in terms of cardiovascular health. Conversely, the greenness accessibility not only contributes to the reduction of perinatal anomalies but also offers a partial amelioration of the adverse impacts from PM_2.5_ and O_3_ exposure. From a policy perspective, our findings underscore the importance of synergistic PM_2.5_-O_3_ control and urban greening for public health benefits.

### Impacts of maternal CVDs on adverse pregnancy outcomes

Maternal primary and pregnancy-induced cardiovascular diseases are significant risk factors for APOs, with an overall HR=1.693 (95% CI: 1.615–1.775, *p*=1.70×10^−26^), indicating pregnant individuals with composite CVDs suffer a 69.3% higher risk to develop adverse gestational outcomes compared to those with normal cardiovascular function. More specifically, pregnancy-induced cardiovascular diseases result in 28.4% higher risks of APOs (HR=1.284, 95% CI: 1.025–1.609, *p*=1.77×10^−2^), whereas primary cardiovascular diseases can lead to a more prominent risk as 37.0% (HR=1.370, 95% CI: 1.304–1.439, *p*=6.81×10^−17^). Fig. 3 presents the association strengths between specific subtypes of maternal cardiovascular disease during pregnancy and APO subsets, revealing in general that pregnancy-induced maternal cardiovascular diseases, particularly hypertension and preeclampsia, exhibit stronger risk associations with various neonatal anomalies compared to primary cardiovascular diseases.

**Fig. 3.**
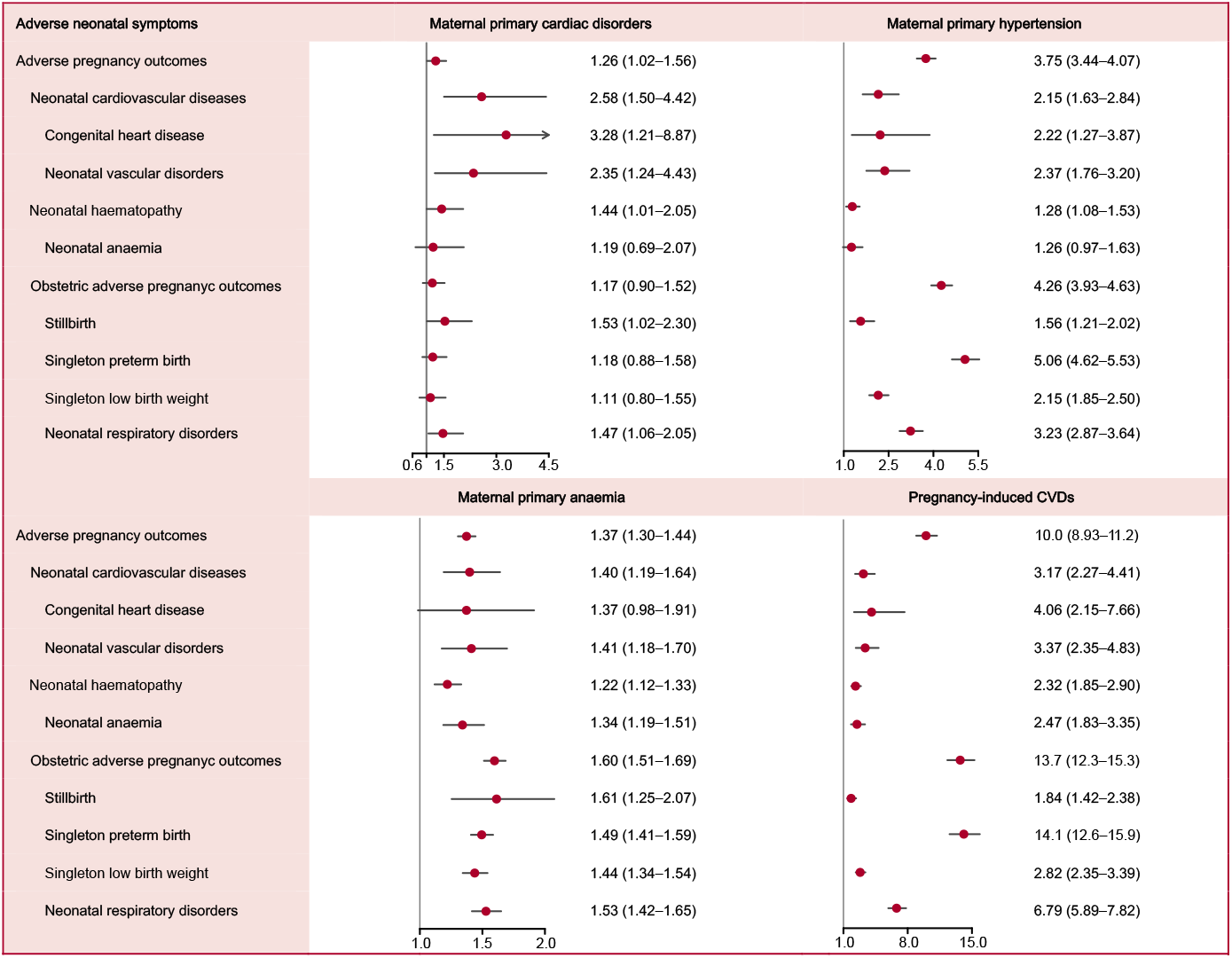
Risk association between maternal cardiovascular and haematological complications and adverse pregnancy outcomes. Risk factors are defined as maternal CVDs, including primary cardiac disorders, primary hypertension, primary anaemia, and pregnancy-induced CVDs. Adverse health outcomes encompass obstetric anomalies (stillbirth, singleton PTB, singleton LBW, and neonatal respiratory disorders), as well as neonatal cardiovascular and haematological disorders. Multiple pregnancies are highly likely to be associated with medically indicated PTB and naturally occurring LBW, and thus are typically excluded from the analysis on investigating obstetric anomalies. Hazard ratios (HR) with 95% confidence intervals (CI) were estimated by Cox regression models having adjusted for covariates for 10-μg/m^3^, 10-ppb, and 0.1-EVI incremental exposure, respectively.

Maternal heart diseases could impact neonatal cardiovascular diseases (Fig. 3). Notably, an observable mother-to-child heritability is evidenced on CHD of HR=2.949 (95% CI: 1.458–5.965, *p*=2.14×10^−3^). It is worth mentioning that a proportion of females born with CHD factually are not recommended for pregnancy. Among 148 expectant mothers with CHD from the ZEBRA maternity cohort, 14 (9.5%) neonates were also diagnosed with CHD, whereas the incidence among the whole cohort was low at 1.2‰. This finding is consistent with previous studies^26^, and the high conditional incidence rate suggests a potential genetic basis for CHD development, although current medical research has yet to fully confirmed the aetiology. As a result, we firmly endorse the implementation of genetic screening among eligible pregnant women with prenatally diagnosed CHD in order to ascertain whether genetic anomalies underlie the congenital defects, since the probability of developing CHD on offspring might be substantially amplified if there are genetic effects, warranting preconception proactive attention in advance.

Maternal vascular diseases exert a greater impact than cardiac defects on the risk of APOs (HR=3.746, 95% CI: 3.445–4.073, *p*=3.71×10^−33^), but maternal blood disorders seem not to significantly affect neonates, excluding anaemia which increases the risk of haematological conditions (HR=1.170, 95% CI: 1.072–1.277, *p*=5.01×10^−4^). Besides neonatal cardiovascular diseases, maternal vascular and blood anomalies also pose risks on other birth defects (Fig. 3). While the majority of neonates (88.5%, 5,842 out of 6,604) did not exhibit congenital cardiovascular anomalies at birth, a significant proportion of premature and low birth weight infants could potentially encounter growth and developmental impairments, culminating in high medical burden and unforeseeable compromise in long-term quality of life^27-29^. This reiterates the significance of extending specialised medical vigilance towards pregnant individuals afflicted by cardiovascular conditions.

Maternal CVDs also modify the risks of APOs from environmental exposures. Pregnant participants with CVDs manifest a 5.2% (4.9–5.5%, *p*=1.85×10^−39^) higher risk regarding the PM_2.5_-APOs association compared to the control group without cardiovascular symptoms, and 2.2% higher (2.1–2.4%, *p*=5.20×10^−29^) on O_3_-APOs risk association. The effect modification of maternal CVDs on greenness-APOs risk association is more pronounced, thereby attenuating the significance of protective effects in certain subtypes of APOs. Fig. 4 provides a comprehensive overview of the intricate effect modifications associated with distinct subtypes of maternal cardiovascular diseases. It demonstrates that maternal CVDs during pregnancy accentuate the susceptibility of predisposed individuals to the adverse impacts of air pollution. For reproductive-age women with primary cardiovascular symptoms, proactive strategies such as minimising exposure to ambient air pollution or actively engaging with green spaces in the preconception period (i.e. at least 6–12 months prior to pregnancy) are advised to attenuate the occurrence of perinatal anomalies.

**Fig. 4.**
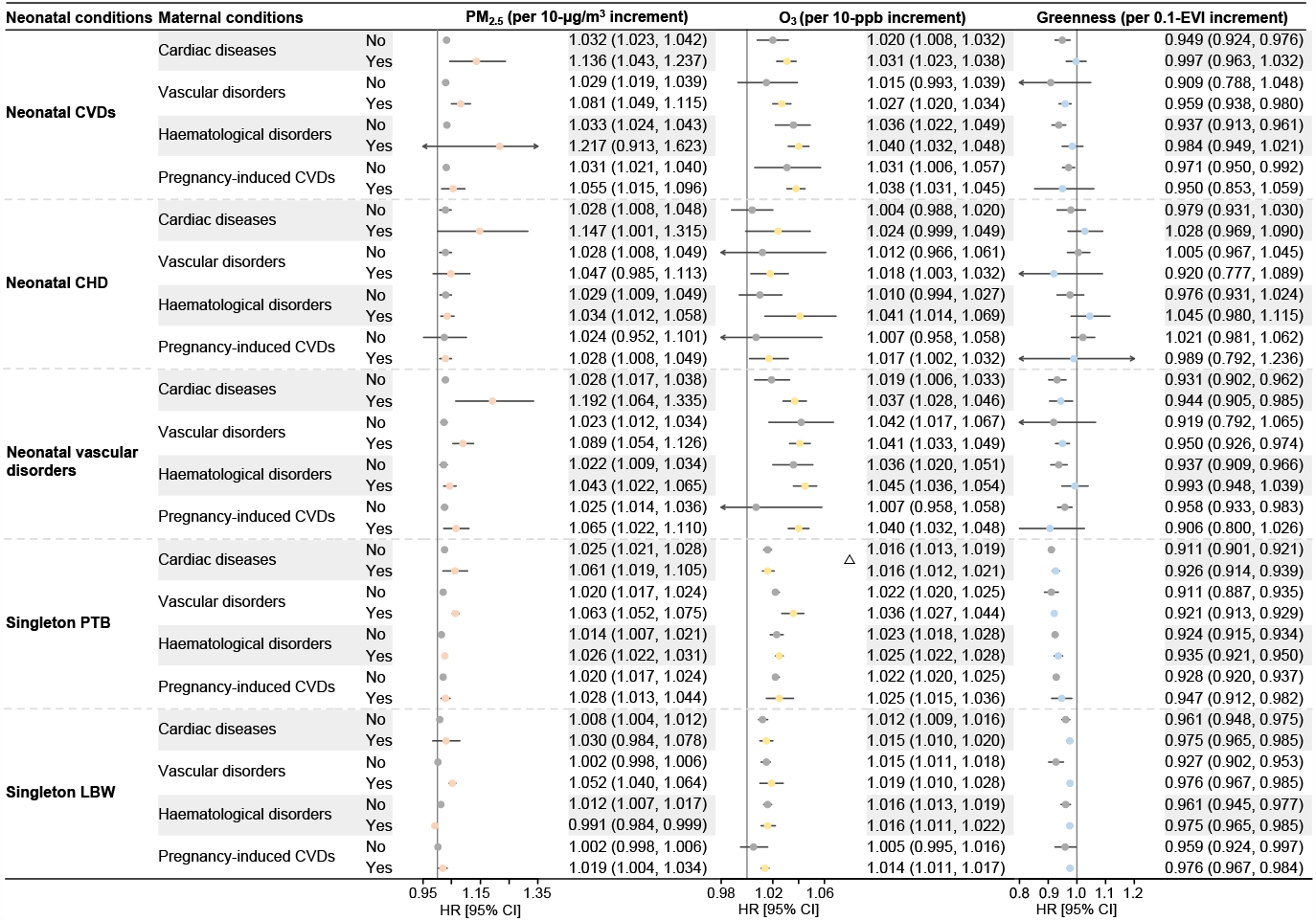
Effect modification of maternal cardiovascular complications on the risk association between environmental exposure and adverse pregnancy outcomes. Four effect modifiers, including maternal primary cardiac diseases, primary vascular disorders, primary haematological disorders, and pregnancy-induced CVDs, were examined. The presentations of estimated hazard ratios (HRs) are defined along three dimensions: neonatal adverse health condition (N), maternal primary and pregnancy-induced diseases (M), and environmental exposure (E), in which context HRs represent the strength of the risk association between E and N, with or without the presence of M. Within the M group, no significant difference at α=0.05 between the two HR levels was indicated by Δ, while the unmarked groups signify that there is a significant difference in risks with or without M.

### Mediation of maternal CVDs between environmental exposure and APOs

From the perspective of a directed acyclic graph, maternal CVDs serve as mediators between environmental exposure and APOs (as illustrated in Extended Data Table 4). Environmental exposures not only pose direct impacts on APOs, but can also exert indirect effects through maternal CVDs. The indirect effects from PM_2.5_, O_3_, and greenness exposure on APOs account for 44.4%, 34.4%, and 23.1% of total effects, respectively, underscoring the significance of mediation by maternal pregnancy-induced CVDs, particularly concerning the risk associations with PM_2.5_ exposure. Environmental exposures still predominantly exert direct effects on APOs, affirming the rationale for prior studies directly investigating the risk associations between environmental exposures and APOs. Indirect effects are most pronounced in PTB and LBW, especially in the impacts on PTB from PM_2.5_ exposure where the indirect effect constitutes over half proportion (50.9%). However, the impact of greenness exposure on APOs is minimally mediated by maternal pregnancy-induced CVDs, particularly in the context of LBW (12.1%) and neonatal CVDs (13.2%). These findings reveal a plausible causality pathway in reproductive epidemiology, where environmental exposure influences maternal cardiovascular function, subsequently impacting pregnancy outcomes. This also suggests the feasibility of reducing the risk of APOs by controlling maternal cardiovascular disorders.

### Ensemble-learning-based risk prediction and self-directed assessment

In virtue of the 60 characteristics collected on the ZEBRA maternity cohort participants together with the 40 environmental exposure tracking metrics, we developed an ensemble deep learning-based prediction model (Extended Data Fig. 4) to forecast the incidence risk of APOs. Since early-stage risk prediction is a continuing pursuit in clinical predictive models (i.e. predicting the occurrence of APOs during the third trimester lacks practical value in timely clinical alert), predictive factors were selected upon the availability before the end of the second trimester. The diagnosis of maternal primary and pregnancy-induced cardiovascular symptoms significantly enhances the risk predictive performances of APOs. Without the cardiovascular diagnoses, sensitivity (i.e. true positive predictions out of all cases) of APOs is limited to 59.7% (false positive rate, FPR=0.8%, area under the ROC curve, AUC=0.899). However, cardiovascular diagnoses reinforce the sensitivity of APOs to 80.8% (FPR=0.4%, AUC=0.956). For the collection of obstetric complications (i.e. PTB and term LBW) and neonatal cardiorespiratory disorders (i.e. CHD, congenital cardiovascular defects, and neonatal asphyxia), the ensemble learning prediction framework achieved accuracies as high as 97.9% (sensitivity=82.8%, FPR=0.1%) and 98.5% (sensitivity=90.6%, FPR<0.1%), respectively. This exemplifies the potential practical value of AI-assisted algorithmic risk prediction models for clinical application in obstetrics and gynaecology, while also reaffirming the paramountcy of monitoring for maternal cardiovascular symptoms during pregnancy.

Nevertheless, the confidentiality of maternal medical records and the restricted interpretability inherent in machine learning algorithms preclude the feasibility of deploying ensemble-learning-based risk prediction frameworks for public utilisation. To cope with this challenge, we developed a self-directed risk assessment questionnaire (Supplementary Table S1) aimed at rapidly estimating APO risk scores by assigning weights to factors easily collectable via self-reporting (Supplementary Table S2), followed by linear summation. Overall, environmental exposures account for approximately 40% of the scoring weights, socioeconomic factors account for 10%, while maternal medical and surgical history account for 50%, among which cardiovascular diseases constitute 40%, comparable to environmental factors (Extended Data Figs. 5–6). While the predictive accuracy of the self-directed form (88.2% for obstetric anomalies, 91.9% for neonatal cardiovascular defects) cannot fully emulate the ensemble-learning-based models, its notable high sensitivities (75.7% for obstetric anomalies, 85.2% for neonatal cardiovascular defects) still retain practical significance, inspiring future in-depth exploration and optimisation.

## DISCUSSION

To the best of our knowledge, this is the first Chinese prospective maternity cohort study comprehensively exploring the risk patterns of maternal and neonatal cardiovascular abnormalities during the perinatal period, providing robust primary epidemiological evidence specifically for the East Asian population. Our current study has four major merits. Firstly, we conducted comprehensive analyses on the risk patterns of maternal and neonatal cardiovascular abnormalities, including various subcategories such as cardiac abnormalities, vascular diseases, and haematological disorders, expanding beyond the conventional focuses merely on stillbirth, PTB, and term LBW^30,31^. Secondly, we assessed the health hazards of multiple environmental factors individually as well as investigated the inter-factor synergistic and antagonistic effects, as literature rarely explores the interactions among risk factors. Thirdly, we quantified the mediating effect of maternal pregnancy-induced cardiovascular complications on the risk association between environmental exposures and APOs, emphasising the importance of protecting the pregnant female as a vulnerable population. Finally, we propose the feasibility of using machine learning frameworks for early-stage APO prediction, based on which the designed self-assessment form demonstrates a methodological innovation.

The APO risk self-assessment form we developed according to epidemiological evidence pioneers a new method for practical public health research, that of translating sophisticated machine-learning-based algorithms into interpretable linear algebra for intuitive weighting of risk factors and convenient risk prediction. We encourage more maternity cohort studies to optimise and upgrade our risk prediction models by reporting potential cross-region heterogeneity, so as to strengthen the population generalisability. When filling the self-assessment form, the environmental exposure levels unavailable for self-report can easily be obtained by cloud matching the residential location and conception dates of the pregnant women with the spatiotemporal resolved environmental records. Therefore, to enable large-scale clinical implementation of APO risk prediction models, the establishment of real-time environmental tracking databases encompassing multiple environmental factors is indispensable.

With an increasing body of epidemiological research (including our current study) revealing the beneficial impacts of greenness exposure on population health, we suggest policy recommendations for nature-based interventions consider: i) enhancing residential greenery, such as the implementation of green roofs and the creation of green streetscapes, particularly in areas heavily affected by air pollution; ii) enhancing the quality of existing parks and gardens, which could involve improving the greenness accessibility, increasing the density of vegetation, and providing outdoor fitness facilities; iii) advising pregnant women with cardiovascular diseases to engage in outdoor nature-based activities as part of their treatment plans; iv) expanding the range of community-led activities within green spaces, which could include initiatives like urban farms and retreat centres; and v) promoting awareness and encouraging participation in wilderness programs, ecotherapy, and the practice of forest bathing^32^.

The self-assessment form designed from the ZEBRA prospective maternity cohort will be continuously calibrated and updated as more participants are recruited and more risk factors are collected, which will be shared on MedRxiv preprint platforms. The intended value of the risk assessment form is enabling the pregnant women to understand her pregnancy risks based on various exposures. Entrusting the complete authority of risk assessment to authoritative institutions may give rise to corruption issues, like healthcare institutions categorising more low-risk pregnant women as high-risk individuals for financial benefits. Empowering pregnant women to be the sword-holders of their own health can result in pre-emptive action which then effectively prevents such unnecessary medical overtreatment.

Our study has several limitations that cannot be addressed at this current stage. Firstly, ZEBRA has not yet collected any genetic information from the cohort participants, which theoretically is the gold standard for distinguishing endogenous differences among populations. Given that genetic markers can often substantially improve the predictive accuracy of disease occurrence^33^, the ZEBRA team plans to collect genetic material of the maternity cohort in the near future. Secondly, we have not measured any internal exposure of organic pollutants such as pesticide residuals, phthalates, and endocrine-disrupting chemicals. However, we have initiated pilot studies to determine the exposure doses of pregnant women and foetuses^34,35^, with the aim of expanding to the entire cohort once the technology matures. Last but not the least, our findings may not be representative of the entire Chinese population due to sparse participant enrolment in Northwest China. Therefore, readers should use our results cautiously, particularly when generalising to broader populations, and hence meta-analyses and heterogeneity tests are strongly recommended when more relevant studies come out. It is optimistically hoped that multi-centre collaborative communities will be established soon to enhance the representativeness and generalisability of epidemiological findings.

## METHODS

### Cohort participant recruitment

From 1 January 2017 to 31 December 2022, ZEBRA maternity cohort enrolled a total of 131,155 parturient women. In addition to the 6,237 participants from the pilot study conducted during 2013–2016^36^, the full cohort now includes a total of 137,392 participants. Although the host institution (Women’s Hospital, Zhejiang University School of Medicine) of ZEBRA is based in Zhejiang Province, it serves as one of the top three flagship maternity and child hospitals in China, attracting expectant mothers from across the nation. Particularly, when pregnant women encounter complex medical conditions during pregnancy and their local healthcare facilities are unable to provide definitive care, they are transferred to Zhejiang, thus joining the ZEBRA maternity cohort. Among the current enrolled cohort participants, 122,440 (89.1%) were from Zhejiang, while 14,952 (10.9%) were from other provinces, making the ZEBRA maternity cohort a nationwide cohort in terms of its scale.

After excluding participants with missing records for more than 20% of the variables (N=16,091), those with gestational ages less than 20 or greater than 44 weeks (N=39), and pregnant women aged under 18 or over 45 years (N=172), our analyses cover 121,090 pregnant women in total (Fig. 1). There were 124,025 live births, with 116,610 singletons, 3,619 pairs of twins, and 59 sets of triplets, after excluding 802 stillbirths.

ZEBRA maternity cohort collects comprehensive sociodemographic and behavioural features from pregnant women residing in Zhejiang Province, China throughout the study period. Trained obstetric nurses conduct questionnaire-based face-to-face interviews to record residential address, household registration, ethnicity, education attainment level, smoking habits (active and second-hand), alcohol history, age at current delivery, gravidity, parity, and date of the last menstrual period. Physical examinations are performed to measure the height and weight of the women before conception and at the end of the second trimester.

Gestational ages are primarily determined through ultrasound examinations conducted in the first or second trimester. In cases where ultrasound records are unavailable, the date of the last menstrual period (LMP) is used as an alternative method to estimate the gestational age. In this study, ultrasound examination was used for 98.4% (N=119,153) of the pregnant women to determine gestational age, while the remaining 1.6% (N=1,937) relied on the date of the last menstrual period.

To ensure data accuracy, the medical history of the pregnant women is cross-referenced with the hospital’s medical records using unique medical IDs. Face-to-face consultations with obstetricians are conducted to verify the referenced information. The cohort profile provides a comprehensive overview of the collected information and the processes involved^24^. Stringent quality control measures are in place, including regular training for healthcare and medical personnel, standardised medical examinations, use of uniform-standard physiological and biochemical measurement equipment, and double validation of questionnaire-based interviews.

### Outcome definition

In our study, we identified primary (referring to the non-pregnancy-cause diseases throughout all ZEBRA-based studies no matter congenital or acquired) and pregnancy-induced (a type of secondary) cardiovascular complications in all enrolled cohort participants. Primary maternal CVDs include primary hypertension (ICD10: I10), pulmonary hypertension (I27), congenital heart diseases (Q20–Q28), heart failure (I30–I45), and cardiac arrhythmias (I47–I49). Haematological disorders (D50–D89) include anaemia (including nutritional anaemia, D50–D53, and haemolytic anaemia, D55–D59), and lymphatic system abnormalities (D76). Pregnancy-induced CVDs include gestational hypertension (O13) and pre-eclampsia (O14). The hierarchical classification and occurrence rates of the studied diseases can be found in Extended Data Fig. 1.

Adverse pregnancy outcomes (APOs) include stillbirth, obstetric anomalies, neonatal cardiovascular diseases, neonatal haematological diseases, and neonatal respiratory disorders. Obstetric anomalies include preterm birth (PTB, O60) and term low birth weight (LBW, P07.1). PTB is defined as gestation less than 37 weeks, and very preterm birth (VPTB) is defined as gestation less than 32 weeks. Term LBW is defined as birth weight less than 2500 grams on the term-birth neonates, while very low birth weight (VLBW) refers to term-birth neonates with birth weight less than 1500 grams. When evaluating risk associations related to obstetric complications, we only considered singleton births.

Neonatal cardiovascular complications mainly include cardiac disorders and pulmonary hypertension. Cardiac abnormalities encompass congenital heart diseases and cardiac arrhythmias, while haematological abnormalities include coagulation defects, purpura, and other haemorrhagic disorders (D65–D69). ZEBRA provides accurate diagnoses for congenital heart diseases in newborns, with subcategories mainly including transposition of great vessels, ventricular septal defects (Q21.0), atrial septal defects (Q21.1), tetralogy of Fallot (Q21.3), patent ductus arteriosus (Q25.0), coarctation of the aorta (Q25.1–Q25.3), and pulmonary stenosis (Q25.5–Q25.6). In this study, we do not further classify maternal congenital heart diseases into subcategories. Neonatal respiratory abnormalities include birth asphyxia (P21) and respiratory distress (P22). The classification and incidence rates of the analysed diseases can be found in Extended Data Fig. 2.

It should be clarified that throughout our present analyses, APOs are defined as all the conditions mentioned above. The cases of PTB and term LBW are counted per pregnancy (equivalent to the number of women with abnormal pregnancy outcomes), as multiple births are often associated with medically induced premature delivery.

### Environmental exposure assessment

In this study, we evaluated maternal exposure to three major environmental factors: ambient PM_2.5_, O_3_, and green space. We tracked the historical concentrations of ambient PM_2.5_ (in daily average) and O_3_ (in daily maximum 8-hour moving average) during the period from 2013 to 2022 using the TAP (Tracking Air Pollution) database^37,38^. The TAP database integrates various data sources, including *in situ* observations, numerical simulations from chemical transport models, satellite-based remote sensing measurements, and land cover information. These data are combined using an ensemble machine learning framework to provide highly accurate daily concentration estimates. The spatial resolution of PM_2.5_ data is 1×1 *km*^2^, while the resolution of O_3_ data is 10×10 *km*^2^. Green spaces are assessed using the Enhanced Vegetation Index (EVI) provided by MODIS Vegetation Index Products^39^. The spatial resolution of EVI data is 0.5×0.5 *km*^2^, and measurements are available every 16 days. Maternal exposures were averaged over an 18-month period, from one year before pregnancy until the end of the second trimester (the 6^th^ month since conception).

We also tracked individual-level exposure to three additional ambient air pollutants, NO_2_, SO_2_, and CO, as covariates for confounding adjustment. These exposure measurements were obtained from other well-established datasets^40-42^. Due to the limitations of historical data availability, the tracking of these air pollutants is only available until December 2020. To overcome this limitation, we extrapolated the pollution concentrations from the corresponding day in 2020 to serve as proxies for the years 2021–2022. Considering the temporal extrapolation and the absence of significant risk associations, the analysis results for these three air pollutants are not presented as the main findings. Furthermore, we accounted for the influence of temperature exposure by controlling for the impact of heat index, which is a humidity-calibrated temperature measure that better captures the perceived sensation of temperature exposure^43,44^. We quantified two heat exposure metrics: daily mean heat index and daily maximum heat index. Daily ambient air temperature data with a resolution of 0.1°×0.1° and humidity data with a resolution of 0.25°×0.25° were retrieved from ECMWF Reanalysis v5 (ERA5) products^45^.

### Statistical analyses

#### Risk association quantification

Extended Cox proportional hazard regression models with time-varying variables were applied to investigate i) the hazard ratios (HR) and 95% confidence intervals (CI) of environmental exposures on maternal pregnancy-induced cardiovascular complications; ii) the HR of environmental exposures on APO and subtypes; and iii) the HR of maternal cardiovascular complications (both primary and pregnancy-induced) on APO and subtypes. Assuming consistent HRs over time, the temporal granularity was delineated on a weekly basis, spanning from the preconception year to the occurrence of the specified health outcomes rather than the conventional gestational age.

The crude regression models solely examine the direct association between the studied risk factor, denoted as *X*_0_, and the occurrence of the target health outcomes (Equation 1). The fully adjusted regression model considers other risk factors and potential confounders (such as other environmental exposure factors, socioeconomic characteristics, medical history, and the sex of neonates, denoted as *X*_1_) (Equation 2). Additionally, interaction terms between the studied pairs of factors are included in the extended model to assess the effect modification (EM) between risk factors (Equation 3).

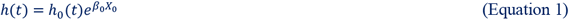

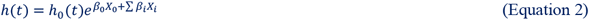

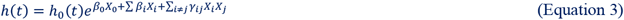

The coefficients *β*_1_ obtained from the Cox regression models accompanied with standard errors (*SE*_1_) are transformed to HR following Equation 4, where Δ*x* represents a unit incremental exposure in the risk factor. For binary variables, Δ*x* is defined as 1, and transformed HRs indicate the increased risk comparing with and without the corresponding risk factor. For environmental exposures, Δ*x* is defined as 10-μg/m^3^, 10-ppb, and 0.1-EVI increments of PM_2.5_, O_3_, and greenness, respectively. The transformed HR signifies the increased risk associated with each 10 μg/m^3^ increment in PM_2.5_ exposure; the interpretations for O_3_ and green space follow the similar way. EM is calculated by Equation 5, which can be interpreted as the alteration of one exposure factor on the association between another environmental exposure and risk of the studied adverse health outcome, under the specified increment of environmental exposure.

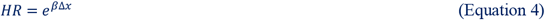

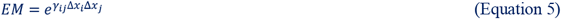

#### Direct and indirect effect evaluation

Given that maternal CVDs are positioned along the causal pathway between environmental exposure and APOs, maternal CVDs are considered mediators from a biostatistical perspective. Multivariate Cox regression models were applied to examine the risk associations quantified in log-transformed HR between i) environmental exposure and APOs having adjusted for all the other covariates except the maternal CVDs, marked as *β*_1_, and ii) environmental exposure and APOs having adjusted for all studied covariates including the maternal CVDs, marked as *β*_2_. *β*_2_ represents the direct effects on APOs from environmental exposure, while *β*_1_ − *β*_2_ indicates the indirect effects. Analysed maternal CVDs as mediators include pregnancy-induced hypertension and preeclampsia, and APOs include PTB, LBW, and neonatal CVDs.

#### Exposure-response trend determination

We employed restricted cubic spline regression with no more than 4 degrees of freedom to investigate the risk association trends of maternal PM_2.5_, O_3_, and greenness exposures on pregnancy-induced cardiovascular diseases (including hypertension), obstetric adverse pregnancy outcomes (including preterm birth and term low birth weight), and neonatal cardiovascular diseases. All covariates considered in the multivariate models are retained. The risk thresholds for the three environmental factors are defined as the 5^th^ percentile of exposure levels among all cohort participants^46^.

#### Inter-group comparison of risk associations

We performed Cox regressions on participants with specific cardiovascular symptoms (including subcategories) and those without specific cardiovascular symptoms to obtain grouped HRs with 95% CIs. Subsequently, we conducted Levene’s tests on the paired log-transformed HRs (i.e. *β*_*i*_ ± *SE*_*i*_) for each group to determine the homogeneity of variances between *SE*_1_ and *SE*_2_, in order to decide whether to use a homoscedastic or heteroscedastic *t*-test. Statistical significance (*p*-value) was defined as *α*<0.05 for two-sided tests; the *t*-test *p*-values (*p*_*hom*_ or *p*_*het*_) are calculated following Equations 6–11, where *n*_1_ and *n*_2_ represent the sample sizes of the two groups; *df* stands for degrees of freedom; *s*_1_ and *s*_2_ for the standard deviations of either group; *s*_*p*_ for the pooled standard deviation; *t*_*hom*_ and *t*_*het*_ for homoscedastic and heteroscedastic *t*-statistics, respectively; *Γ* for gamma distribution; *p*_*i*_ for *p*-values of homoscedastic and heteroscedastic *t*-tests, respectively.

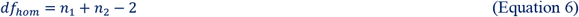

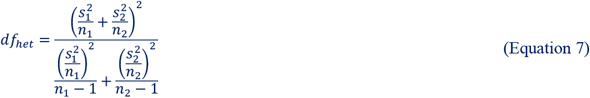

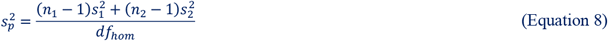

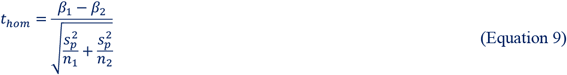

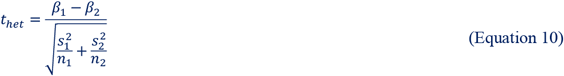

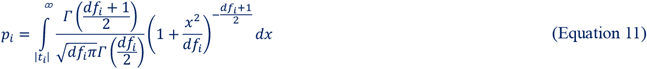

All statistical analyses in this study were performed in Stata 17. The prediction model was performed via the “*scikit-learn*” (version 1.2.0) and “*xgboost*” package (version 1.6.2) in Python 3.8.0. Computations were supported by JASMIN supercomputer.

### Ensemble machine learning algorithm for risk prediction

We integrated 10 socioeconomic features, 30 environmental exposure indicators, 10 items for pregnancy-relevant maternal cardiovascular diagnoses, 21 items for obstetric disease diagnoses, and 29 items of other aspects of medical history to predict the risks of obstetric APOs and neonatal cardiovascular diseases by virtue of ensemble learning. Initially, we employed 6 classical base learners, as i) linear logistic regression classifier, ii) decision tree classifier, iii) random forest classifier, iv) extra-trees classifier, v) bootstrap aggregating (bagging) classifier, and vi) gradient boosting classifier, to construct the supervised training models and generate predicted probability scores of the studied adverse health outcomes. Subsequently, we used a fully connected multi-layer perceptron classifier to integrate the original predictive factors and the risk probability scores obtained from the base learners to build a final predictive model for the occurrence of the two categories of APOs. The multi-layer perceptron classifier consists of a fully connected artificial neural network with 5 hidden layers, each composed of 256, 256, 128, 64, and 32 nodes, respectively (refer to Extended Data Fig. 4 for the algorithm structure). The hyperparameters (number of hidden layers and nodes) of the multi-layer perceptron are determined by learning curves.

We evaluated the predictive performance of the final multi-layer perceptron classifier by randomly selecting 80% of the samples for model training and performed 10-fold cross-validation tests; the remaining 20% of the samples (N=24,218) were used for external validation of the model. Evaluation also included assessing the sensitivity, false positive rate, and the overall power of the prediction model (area under the ROC curve, AUC).

### Design of self-directed APO risk assessment form

We utilised ensemble learning algorithms to forecast the probability of risk events transpiring, employing the prediction score as the dependent variable. We consider all discretised risk factors as independent variables and ascertain the weighting coefficients through linear regression. Continuous variables were discretised using quartiles (e.g. environmental exposure levels) or predetermined segmenting criteria (e.g. BMI). The procedure for determining the weights of the risk factors can be outlined as follows:

1. Perform a linear regression with intercept *β* on the target score *P* against the discrete risk factors ***X***_*D*_, generating an initial weight matrix ***A***_0_, in terms of *P =* ***A***_0_***X***_*D*_ + *β*.
2. For risk factors with negative weights, reverse the sequence of the corresponding discrete labels 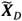, and transform the weight matrix ***A***_0_ into a non-negative matrix, 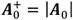.
3. Define 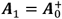, where subscript indicates the round of iteration. **Iteration start:**
  - Compute the weighted score excluding the intercept: 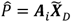.
  - Select the case-control overlapping samples, based on 95% interval (2.5–97.5^th^ percentile) of the score distribution for cases and controls.
  - Perform a linear regression with intercept on sample set, generate a new weight matrix, ***A***^′^, for the current iteration.
  - Adjust the weight matrix: 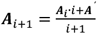
  - Check if either i) |***A***_*i*+1_ − ***A***_*i*_| <0.001, or ii) the number of overlap samples is less than 5% of the total number of cases.
  **Iteration ends**.
4. Calculate the sum of the maximum weights for each factor under the adjusted weight matrix after iteration, ***A***_*i*+1_, and rescale it to 100, yielding the final weights, ***A***_*s*_.
5. Compute the fully optimised risk scores for all pregnant women in the cohort with rescaled weights: 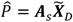. Define the median score of the overlap interval for cases and controls as the risk forecast threshold.
6. Complete the tabulation (see Supplementary Table S1 for a sample table).

We conducted external validation for the self-directed APO risk prediction form on 5,217 pregnant women recruited between 1 January and 31 May 2023. The de-identified questionnaires of two pregnant women with the highest predicted risk scores of obstetric APOs and neonatal cardiovascular defects are enclosed in Supplementary Tables S3–S4 as samples.

### Sensitive analysis

We conducted multiple sensitivity analyses to ensure the robustness of our results. These analyses include: 1) randomly selecting 50% of the original samples for five times of statistical analysis (Supplementary Tables S5–S6); 2) using different combinations of environmental exposures to adjust for potential confounders (Supplementary Table S7); and 3) dividing the whole studied population into two subgroups, within Zhejiang and outside Zhejiang Province, so as to test the inter-group heterogeneity of the estimated risk associations (Supplementary Table S8).

### Ethics Approval

The current cohort-based study was approved by the ethics committee of the Women’s Hospital, Zhejiang University School of Medicine (IRB-20220189-R) and we obtained written informed consent from all cohort participants upon enrolment. As an original investigation providing first-hand epidemiological evidence, Strengthening the Reporting of Observational Studies in Epidemiology (STROBE) guidelines for result reporting (see in Supplementary Table S9) was strictly followed.

## Data Availability

The information of the ZEBRA maternity cohort participants for privacy protection it is not disclosed for public use. Researchers interested in accessing the data are encouraged to contact the principal investigators with a brief research proposal. Access to the data will be granted after approval by the ZEBRA committee and the Health Commission of Zhejiang Province. The codes for analysis can be shared upon request.

## EXTENDED DATA

**Extended Data Fig. 1.**
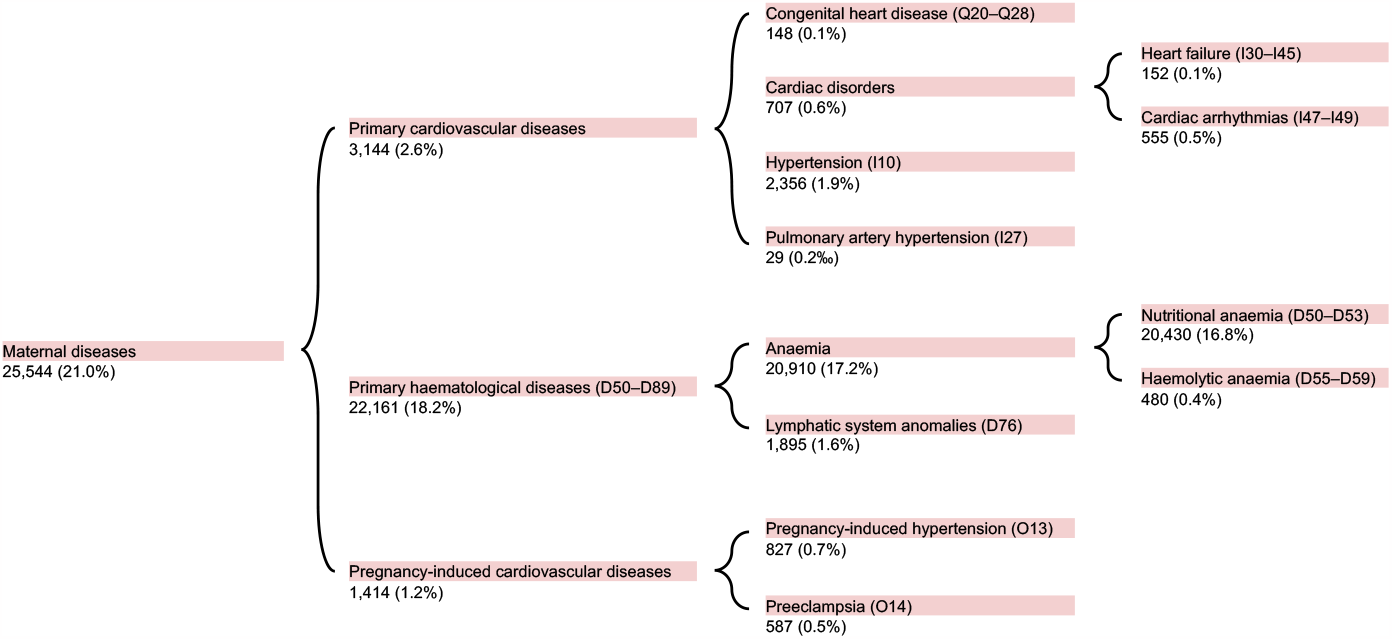
Incidence of adverse maternal health outcomes by hierarchical categorisation. Studied maternal diseases encompass three major categories: primary CVDs, primary haematological diseases, and pregnancy-induced CVDs. The diagnosis of specific diseases corresponds to the International Classification of Diseases 10^th^ Revision (ICD-10) coding system. The number of cases and incidence rates for each hierarchical level of disease are indicated beneath the disease labels.

**Extended Data Fig. 2.**
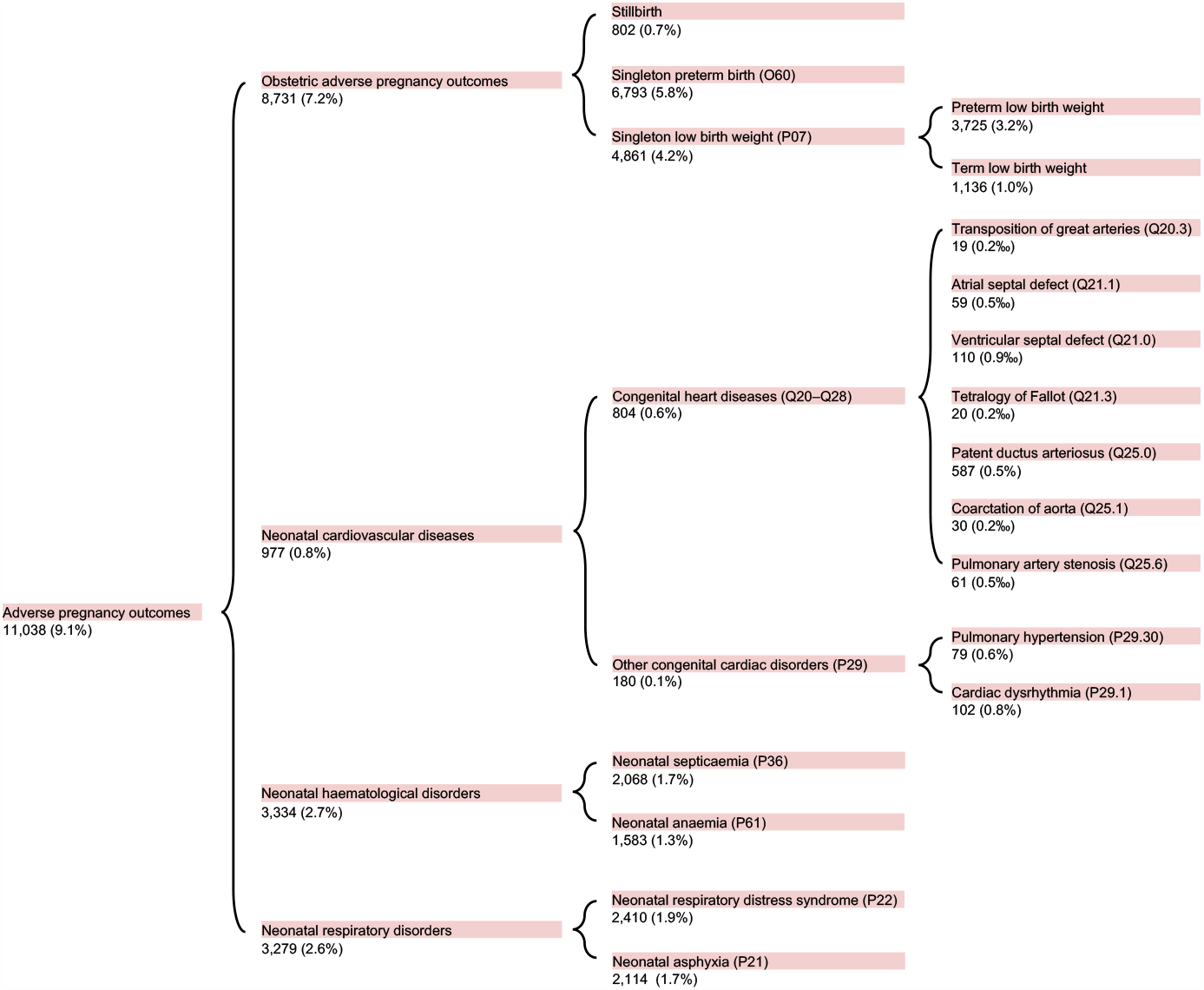
Incidence of adverse pregnancy outcomes by hierarchical categorisation. Adverse pregnancy outcomes encompass four aspects: obstetric anomalies, neonatal CVDs, neonatal haematological diseases, and neonatal respiratory disorders. The figure configuration follows the layout of Extended Data Fig. 1.

**Extended Data Fig. 3.**
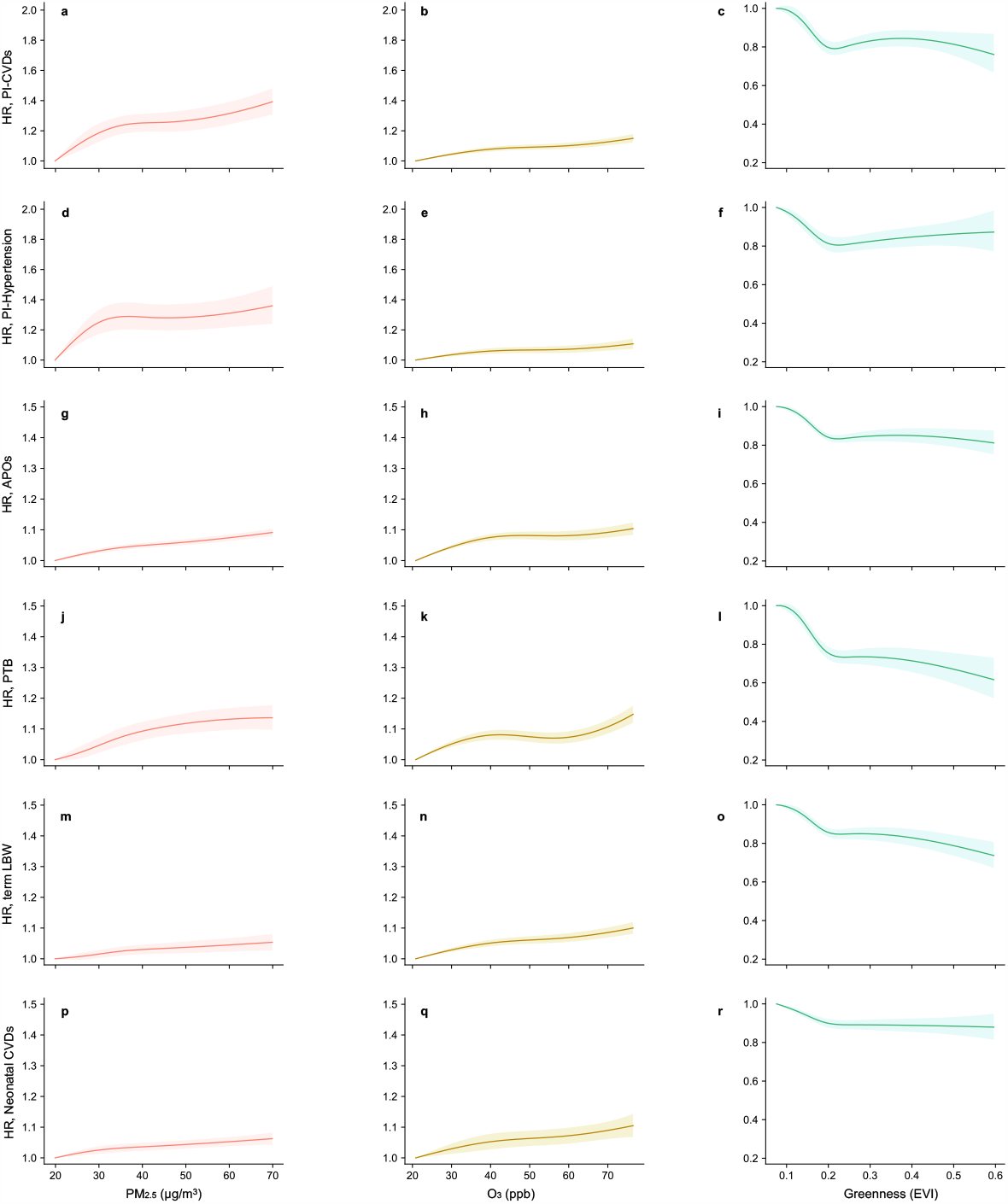
Risk association curves of environmental exposures on adverse maternal pregnancy-induced cardiovascular complications and adverse pregnancy outcomes. Restricted cubic spline regression models with no more than 4 degrees of freedom are applied to investigate the curved risk association trends (quantified in hazard ratio, HR) between maternal PM_2.5_, O_3_, greenness exposure (presented in columns) and adverse health outcomes. **a-c**, pregnancy-induced cardiovascular diseases; **d-f**, pregnancy-induced hypertension; **g-i**, adverse pregnancy outcomes, **j-l**, preterm birth, **m-o**, term low birth weight, **p-r**, neonatal cardiovascular diseases. Thresholds are defined as the lowest 5^th^ percentiles of the exposure levels among the studied cohort participants, as 19.8 μg/m^3^, 20.9 ppb, and 0.076 EVI for PM_2.5_, O_3_, and greenness, respectively. The trend observation ranges are determined by 1^st^-99^th^ percentiles, as 16.4–76.4 μg/m^3^, 16.1–76.2 ppb, and 0.044–0.542 EVI for PM_2.5_, O_3_, and greenness, respectively.

**Extended Data Fig. 4.**
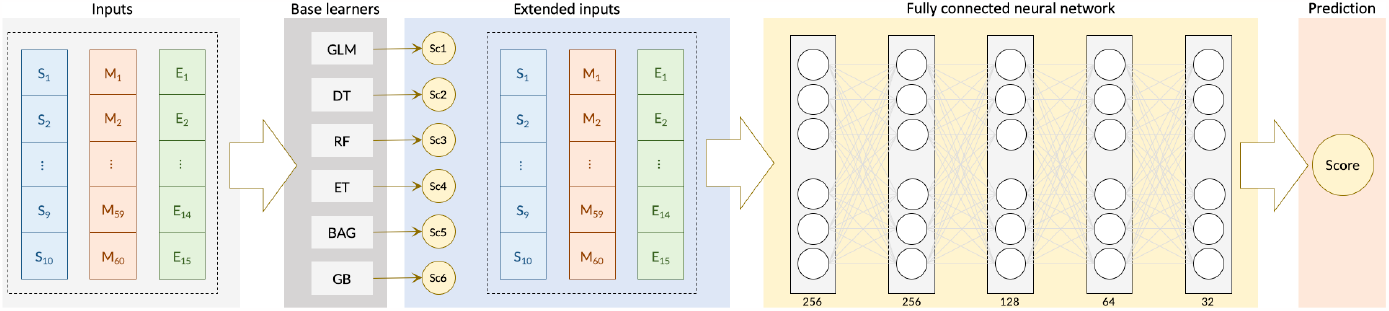
Schematic diagram of ensemble-learning-based risk prediction algorithm framework for adverse pregnancy outcomes. The prediction targets of the algorithm are i) total obstetric anomalies and ii) total neonatal CVDs, without distinguishing any subsets. Initially, the algorithm takes 10 socioeconomic features (denoted as S), 60 medical diagnostic records (M), and 15 environmental exposure factors (E) as input layers. It conducts first-stage risk prediction using six base learners: generalised linear model (GLM), decision tree classifier (DT), random forest classifier (RF), extra-tree classifier (ET), bootstrap aggregating classifier (BAG), and gradient boosting classifier (GB). The predicted risk scores (Sc) obtained in the first-stage algorithm, together with the raw initial inputs, serve as the new input layers for the second-stage fully connected neural network (FCNN). The FCNN comprises five hidden layers, each consisting of 256, 256, 128, 64, and 32 nodes, respectively. The outputs of the second-stage algorithm ultimately represent the predicted occurrence probabilities for the two designed prediction targets.

**Extended Data Fig. 5.**
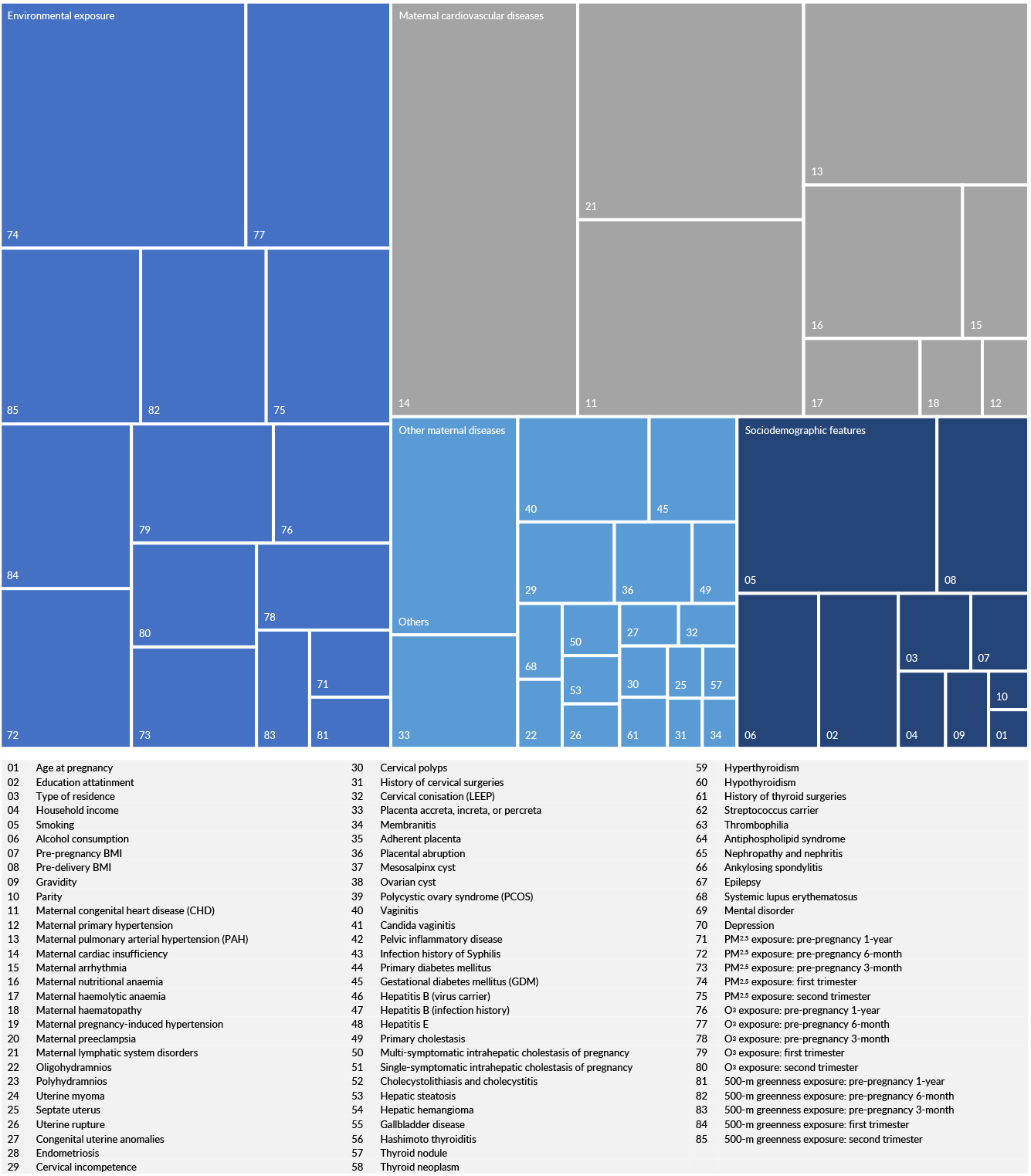
Treemap chart of weighted risk scores of obstetric adverse pregnancy outcomes. The treemap illustrates the weights of the 85 risk factors used for prediction via the self-assessment form. These risk factors are categorised into four groups: sociodemographic features, environmental exposure, maternal CVDs, and other maternal diseases, comprising 12.6%, 38.0%, 34.5%, and 14.9% of the total weight, respectively. In clinical practice, pregnant women who develop pregnancy-induced hypertension or even preeclampsia are almost inevitably subjected to caesarean section, leading to medically indicated preterm birth. Therefore, maternal pregnancy-induced hypertension and preeclampsia were excluded in risk weighting.

**Extended Data Fig. 6.**
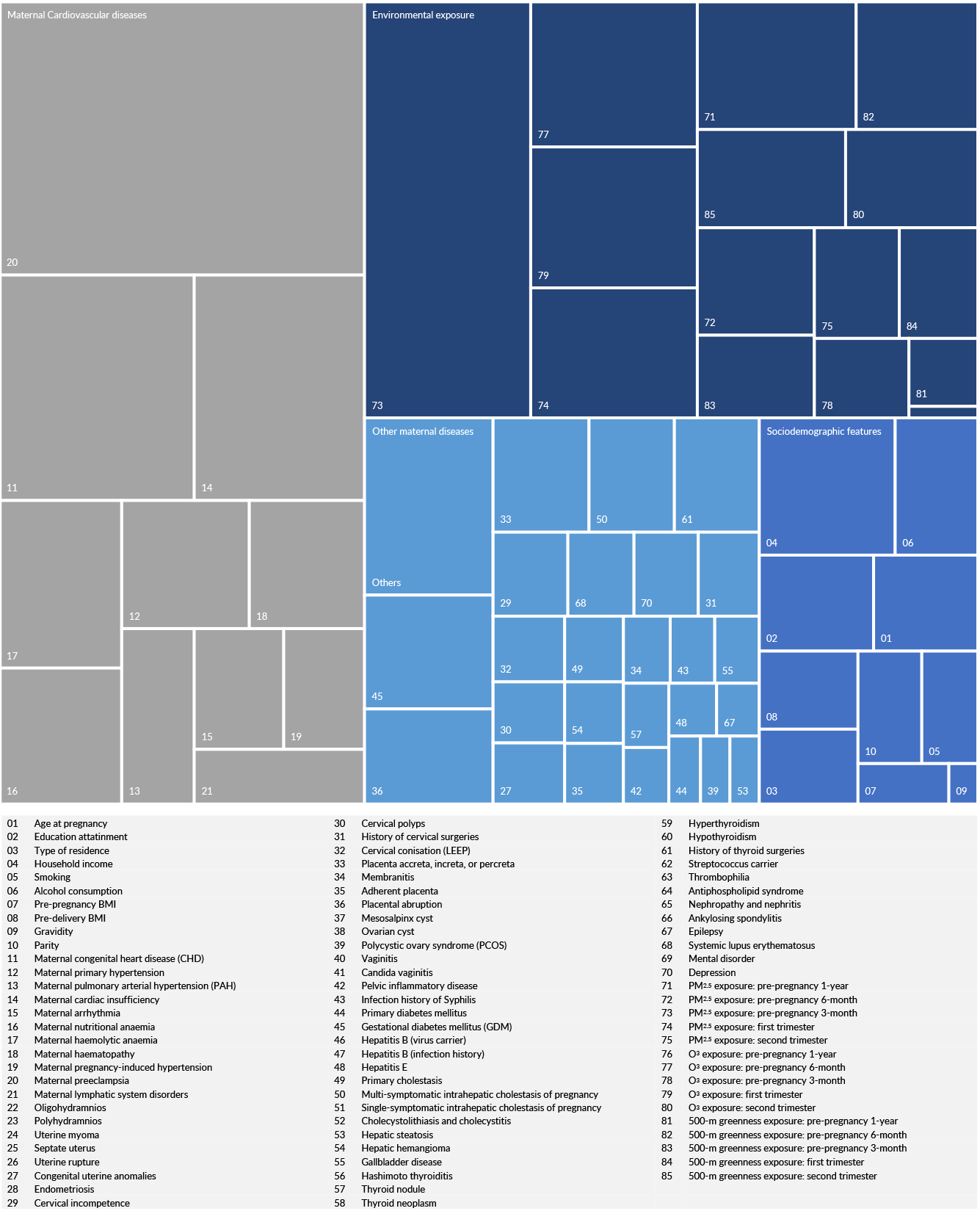
Treemap chart of weighted risk scores of neonatal cardiovascular diseases. The layout of the figure follows the configuration presented in Extended Data Fig. 5. The four groups of risk factors: sociodemographic features, environmental exposure, maternal CVDs, and other maternal diseases, account for weights of 10.7%, 32.5%, 37.3%, and 19.5%, respectively.

**Extended Data Table 1.**
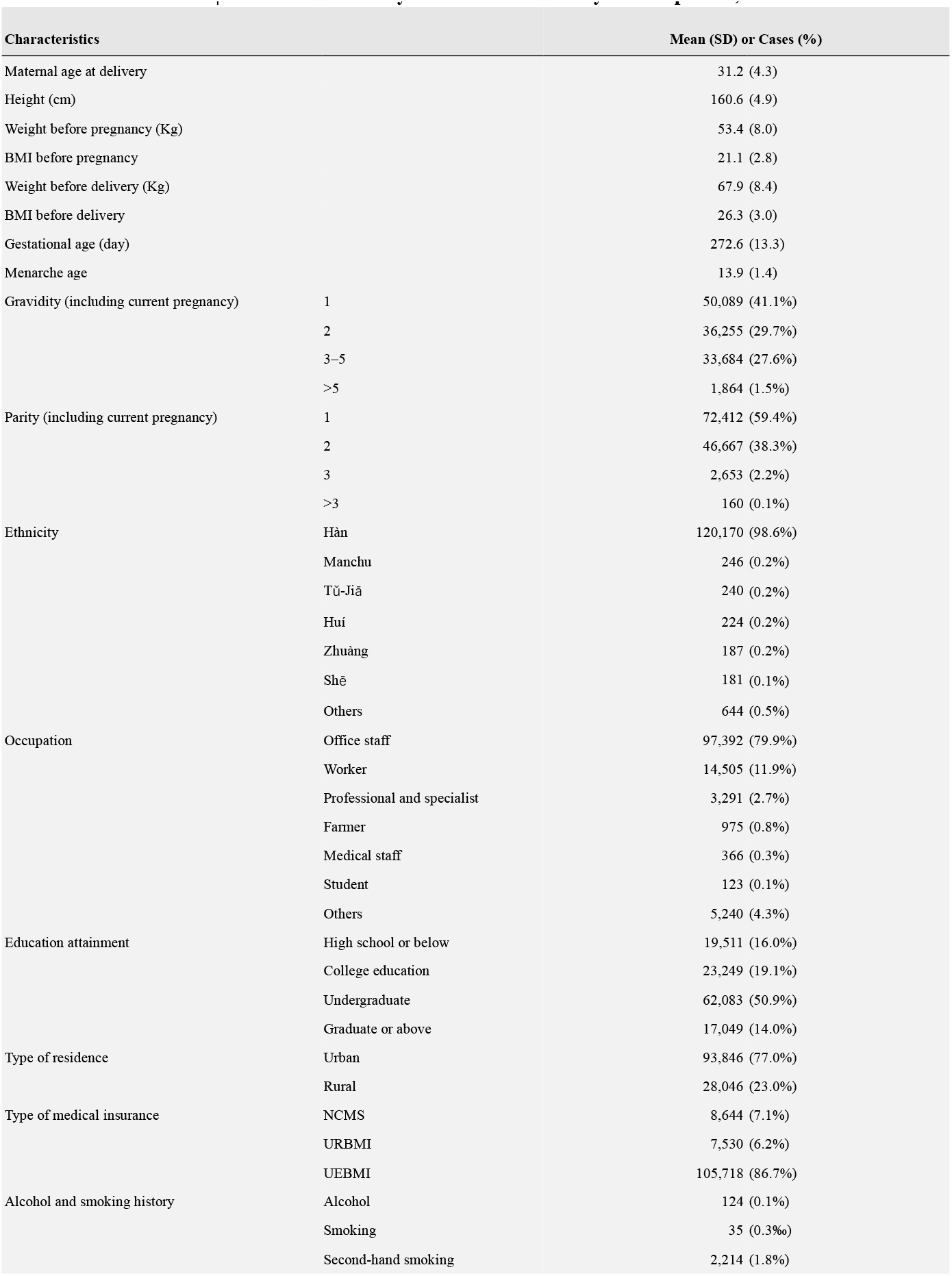
Statistical summary of ZEBRA maternity cohort profile, 2013–2022.

**Extended Data Table 2.** Statistical summary of medical history.

| History of diseases and medical treatments | Cases (%) |
| --- | --- |
| Uterine fibroids | 7,951 (6.5%) |
| Septate uterus | 613 (0.5%) |
| Uterine rupture | 474 (0.4%) |
| Congenital uterine malformation | 293 (0.2%) |
| Cervical incompetence | 1,107 (0.9%) |
| History of cervical surgery | 687 (0.6%) |
| Cervical polyps | 427 (0.4%) |
| Cervical conisation (LEEP) | 314 (0.3%) |
| Membranitis | 527 (0.4%) |
| Placenta accreta, increta, or percreta | 825 (0.7%) |
| Morbidly adherent placenta | 3,369 (2.8%) |
| Placental abruption | 2,675 (2.2%) |
| Mesosalpinx cyst | 2,758 (2.3%) |
| Ovarian cyst | 2,136 (1.8%) |
| Polycystic ovary syndrome (PCOS) | 356 (0.3%) |
| Vaginitis | 552 (0.5%) |
| Pelvic inflammatory disease (PID) | 455 (0.4%) |
| Systemic lupus erythematosus | 71 (0.1%) |
| Syphilis (infection history) | 423 (0.3%) |
| Gestational diabetes mellitus | 290 (0.2%) |
| Antiphospholipid syndrome (APS) | 902 (0.7%) |
| Hepatitis B (virus carrier) | 6,825 (5.6%) |
| Hepatitis B (infection history) | 5,349 (4.4%) |
| Hepatic haemangioma | 64 (0.1%) |
| Gallbladder diseases | 386 (0.3%) |
| Single-symptomatic intrahepatic cholestasis of pregnancy | 2,864 (2.3%) |
| Multi-symptomatic intrahepatic cholestasis of pregnancy | 805 (0.7%) |
| Cholecystolithiasis and cholecystitis | 313 (0.3%) |
| Hepatic steatosis (HS) | 61 (0.1%) |
| Other liver diseases or anomalies | 1,932 (1.6%) |
| Hashimoto thyroiditis | 1,043 (0.9%) |
| Thyroid cancer | 720 (0.6%) |
| Hyperthyroidism | 231 (0.2%) |
| Hypothyroidism | 406 (0.3%) |
| History of thyroid surgery | 402 (0.3%) |
| Kidney diseases | 255 (0.2%) |
| Endometriosis | 815 (0.7%) |
| Thrombophilia | 2,031 (1.7%) |
| Ankylosing spondylitis | 160 (0.1%) |
| Epilepsy | 88 (0.1%) |
| Mental disorders | 82 (0.1%) |
| Depression | 187 (0.2%) |

**Extended Data Table 3.** Sensitivity analyses on risk associations by adjusting for combinations of confounders.

| Confounder Adjustment | PM<sub>2.5</sub> (10 µg/m<sup>3</sup>) | O<sub>3</sub> (10 ppb) | Greenness (0.1 EVI) |
| --- | --- | --- | --- |
| **Maternal pregnancy-induced cardiovascular diseases** |  |  |  |
| Crude HR | 1.053 (1.046–1.060) | 1.006 (1.002–1.011) | 0.946 (0.931–0.962) |
| Combo 1: Physiological and lifestyle characteristics, including age at pregnancy, BMI, ethnicity, alcohol and smoking history. | 1.050 (1.043–1.058) | 1.008 (1.003–1.013) | 0.947 (0.931–0.963) |
| Combo 2: Combo 1 + socioeconomic status, including urban/rural residence, medical insurance, occupation, education attainment. | 1.054 (1.047–1.062) | 1.008 (1.003–1.013) | 0.929 (0.914–0.945) |
| Combo 3: Combo 2 + obstetric and relevant surgical history, including gravidity, parity, history of cervical, hysteroscopic and placental surgeries. | 1.057 (1.050–1.064) | 1.008 (1.003–1.013) | 0.930 (0.915–0.946) |
| Combo 4: Combo 3 + non-cardiovascular medical history, including liver and gallbladder diseases, thyroid diseases, metabolic disorders, immune system diseases, mental disorders, etc. | 1.057 (1.050–1.064) | 1.008 (1.003–1.013) | 0.931 (0.916–0.947) |
| Combo 5: Combo 4 + cardiovascular and haematological medical history. | 1.055 (1.048–1.062) | 1.009 (1.005–1.014) | 0.931 (0.916–0.947) |
| Combo 6: Combo 5 + multiple studied environmental exposures, as PM<sub>2.5</sub>, O<sub>3</sub>, and greenness. | 1.070 (1.061–1.079) | 1.029 (1.024–1.035) | 0.968 (0.951–0.985) |
| Combo 7: Combo 6 + additional environmental exposures, as NO<sub>2</sub>, SO<sub>2</sub>, CO, temperature and humidity. | 1.073 (1.064–1.082) | 1.027 (1.022–1.033) | 0.964 (0.948–0.982) |
| **Adverse pregnancy outcomes** |  |  |  |
| Crude HR | 1.013 (1.010–1.016) | 1.010 (1.008–1.012) | 0.974 (0.968–0.980) |
| Combo 1: Physiological and lifestyle characteristics, including age at pregnancy, BMI, ethnicity, alcohol and smoking history. | 1.012 (1.009–1.015) | 1.010 (1.009–1.012) | 0.977 (0.971–0.983) |
| Combo 2: Combo 1 + socioeconomic status, including urban/rural residence, medical insurance, occupation, education attainment. | 1.015 (1.013–1.018) | 1.011 (1.009–1.013) | 0.955 (0.949–0.961) |
| Combo 3: Combo 2 + obstetric and relevant surgical history, including gravidity, parity, history of cervical, hysteroscopic and placental surgeries. | 1.018 (1.015–1.021) | 1.010 (1.008–1.012) | 0.951 (0.945–0.957) |
| Combo 4: Combo 3 + non-cardiovascular medical history, including liver and gallbladder diseases, thyroid diseases, metabolic disorders, immune system diseases, mental disorders, etc. | 1.018 (1.015–1.021) | 1.010 (1.008–1.012) | 0.952 (0.946–0.958) |
| Combo 5: Combo 4 + cardiovascular and haematological medical history. | 1.016 (1.013–1.018) | 1.011 (1.009–1.013) | 0.951 (0.945–0.957) |
| Combo 6: Combo 5 + multiple studied environmental exposures, as PM<sub>2.5</sub>, O<sub>3</sub>, and greenness. | 1.019 (1.016–1.023) | 1.016 (1.014–1.019) | 0.960 (0.954–0.967) |
| Combo 7: Combo 6 + additional environmental exposures, as NO<sub>2</sub>, SO<sub>2</sub>, CO, temperature and humidity. | 1.015 (1.012–1.019) | 1.017 (1.015–1.019) | 0.961 (0.954–0.967) |

**Extended Data Table 4.**
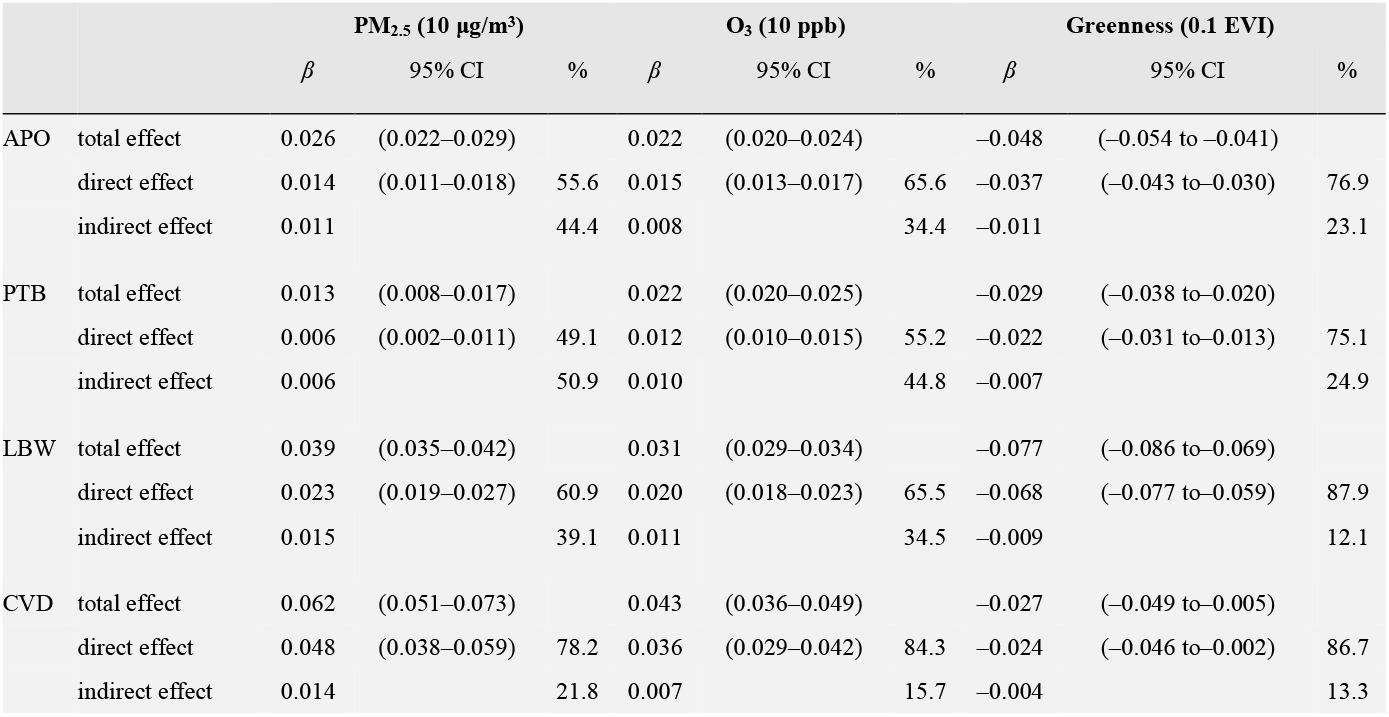
Direct and indirect effects of maternal pregnancy-induced cardiovascular diseases as mediator between environmental exposure and adverse pregnancy outcomes. Total adverse pregnancy outcomes (APO) together with three subsets, preterm birth (PTB), low birth weight (LBW), and neonatal cardiovascular diseases (CVD) are assessed for mediation effects via maternal pregnancy-induced CVDs. Environmental exposures include PM_2.5_, O_3_ and greenness, quantified in 10-μg/m^3^, 10-ppb, and 0.1-EVI increment, respectively. Effects are quantified in *β*, log-transformed hazard ratio (HR), estimated from Cox proportional hazard regression models with 95% confidence intervals (CI). Proportions of direct and indirect effects are estimated in percentages (%).

## Acknowledgements

This study is funded by the Zhejiang Province Health Innovative Talent Project (A0466), International Cooperation Seed Program of Women’s Hospital, Zhejiang University (GH2022B008-01), Key Projects of the Science and Technology Co-construction by National Administration of Traditional Chinese Medicine and Zhejiang Provincial Administration of Traditional Chinese Medicine (GZY-ZJ-KJ-23082), Australian Research Council (DP210102076), and Australian National Health and Medical Research Council (APP2000581). Jason Sun also thanks generous support from the U.S. Fulbright Program. Yuming Guo is supported by a Career Development Fellowship of the Australian National Health and Medical Research Council (APP1163693). Special appreciations to Professor Weiguo Lu, the director of the Key Laboratory of Women’s Reproductive Health, for his comprehensive supports in establishing and coordinating the maternity cohort.

## Author contributions

J.S., Y.G., and X.B. conceptualised and designed the study. W.X. and X.B. were responsible for the administration of the maternity cohort. H.T., H.Z., Q.X., Y.T., and other non-academic members from the ZEBRA collaborative group collected, censored, and pre-processed medical records and associated data for analysis. J.S. performed analyses and wrote the manuscript with comprehensive supports from all authors. K.R.D., K.T., E.X.L., and L.P.S. supervised the study design, reviewed and edited the manuscript.

## Additional information

Supplementary Materials include 6 long-list supplementary contents and 4 tables in 55 pages.

## SUPPLEMENTARY TABLES

Supplementary Table S1 | Self-directed risk assessment questionnaire for early-stage forecasting of obstetric adverse pregnancy outcomes and neonatal congenital cardiovascular diseases.

Supplementary Table S2 | Scoring for risk factors of self-directed questionnaire for early-stage risk forecasting of obstetric adverse pregnancy outcomes and neonatal congenital cardiovascular diseases.

Supplementary Table S3 | A sample filled self-directed questionnaire with the highest risk score of obstetric ad-verse pregnancy outcomes.

Supplementary Table S4 | A sample filled self-directed questionnaire with the highest risk score of neonatal cardiovascular diseases.

Supplementary Table S5 | Sensitivity analysis: Fully adjusted risk associations between environmental exposure and adverse health outcomes by bootstrap resampling.

Supplementary Table S6 | Sensitivity analysis: Fully adjusted risk associations between maternal cardiovascular diseases (primary and pregnancy-induced) and adverse pregnancy outcomes (obstetric and neonatal diseases) by bootstrap resampling.

Supplementary Table S7 | Sensitivity analysis: Risk associations between environmental exposure and adverse health outcomes by different combination of multi-factor adjustment.

Supplementary Table S8 | Sensitivity analysis: Robustness of estimated risk associations at Zhejiang Provincial scale and China nationwide scale.

Supplementary Table S9 | STROBE checklist: Checklist of items that should be included in reports of observational studies.

Supplementary Table S10 | ZEBRA Collaborative Group full roster.

