## Supplementary Materials for "Maternal cardiovascular and haematological complications alter the risk associations between environmental exposure and adverse pregnancy outcomes"

*Supplementary Materials include 6 long-list supplementary contents and 4 tables in 55 pages.*

### Supplementary Table S1 | Self-directed risk assessment questionnaire for early-stage forecasting of obstetric adverse pregnancy outcomes and neonatal congenital cardiovascular diseases.

This is a deformatted self-directed risk assessment questionnaire presented in the form of a linear table for the sake of ease of formatting in Supplementary Materials. The questionnaire in simplified Chinese language version was put into clinical application in Zhejiang province since 1 January 2023, and is identified with an internal beta version series number as beta 1.0: *Self-directed risk assessment questionnaire for early-stage forecasting of obstetric adverse pregnancy outcomes and neonatal congenital cardiovascular diseases (1.0 Beta)*. Since the self-administered questionnaire involves diseases specific to the mid-to-late pregnancy period (such as intrahepatic cholestasis of pregnancy and preeclampsia), the target participants for this questionnaire are the pregnant women in 5-6$^{th}$ month of pregnancy. The self-administered questionnaire for clinical use requires subjects to provide identity-related information, which has been omitted in this research paper, retaining only the ZEBRA-exclusive ID used for case tracking (distinguished from the any official ID like national identification number or social security number of any pregnant woman). The risk factors within each category are not arranged in order of risk score to prevent participants from detecting the degree of risk posed by each factor, which could potentially interfere with the reliability of self-reporting.

#### A. Residence information for the current pregnancy

*Section A is not directly involved for risk scoring*

**A-1. Start date of the current pregnancy (preferred diagnosis from ultrasound examination):**

________________________________

**A-2. Did you change your residential address between one year before conception and the day of delivery?**

○ Yes

○ No

If “No”, please provide:

**A-2.1 Permanent address (Province, City, and Street):**

________________________________

Permanent address longitude (not filled by the participants, but geocoded from address texts):

________________________________

Permanent address latitude:

________________________________

If “Yes”, please provide:

**A-2.2.1. Address 1 (Province, City, and Street):**

________________________________

Address 1 longitude:

________________________________

Address 1 latitude:

________________________________________

**A-2.2.2. Residence period at Address 1 (precise to dates):**

________________________________________

**A-2.2.3. Address 2 (Province, City, and Street):**

________________________________________

Address 2 longitude:

________________________________________

Address 2 latitude:

________________________________________

**A-2.2.4. Residence period at Address 2 (precise to dates):**

________________________________________

**A-2.2.5. Address 3 (Province, City, and Street):**

________________________________________

Address 3 longitude:

________________________________________

Address 3 latitude:

________________________________________

**A-2.2.6. Residence period at Address 3 (precise to dates):**

________________________________________

### B. Basic socioeconomic characteristics

*10 risk scoring features*

**B-1. Age at the current pregnancy:**

○ ≤29
○ 29–39
○ >39

**B-2. Hukou category (household registration type):**

○ Rural
○ Rural-to-urban migration
○ Urban

**B-3. Highest educational attainment level:**
○ Elementary school or below
○ High school or technical school
○ Undergraduate or college
○ Postgraduate or higher

**B-4. Household income level:**
○ ≤5,000
○ 5,000–20,000
○ 20,000–50,000
○ >50,000

**B-5. Smoking habit:**
○ No
○ Passive smoking (current or history)
○ Active smoking (current or history)

**B-6. Alcohol consumption habit:**
○ No
○ Yes (current or history)

**B-7. Height (in metre):**

___________________________________

**B-8. Pre-pregnancy Weight (in Kg):**

___________________________________

**B-9. Weight-gain upon delivery (in Kg):**

___________________________________

**B-10. Pre-pregnancy BMI (automatically calculated):**
○ ≤18.5
○ 18.5–23.9
○ >23.9

**B-11. BMI upon delivery (automatically calculated):**
○ ≤29.8
○ 29.8–32.8
○ >32.8

**B-12. Parity of the current pregnancy:**
○ 1
○ 2–3

○ >3

**B-13. Gravidity of the current pregnancy:**
○ 1
○ 2–3
○ >3

### C. Cardiovascular and haematological conditions diagnosed at the current pregnancy

*11 risk scoring indicators*

**C-1. Nutritional anaemia:**
○ Yes
○ No

**C-2. Haemolytic anaemia:**
○ Yes
○ No

**C-3. Congenital heart disease (CHD):**
○ Yes
○ No

**C-4. Pulmonary arterial hypertension (PAH):**
○ Yes
○ No

**C-5. Primary non-pregnancy-induced hypertension:**
○ Yes
○ No

**C-6. Pregnancy-induced hypertension:**
○ Yes
○ No

**C-7. Preeclampsia:**
○ Yes
○ No

**C-8. Cardiac insufficiency:**
○ Yes
○ No

**C-9. Arrhythmia:**

○ Yes
○ No

**C-10. Lymphatic system diseases:**
○ Yes
○ No

**C-11. Non-anaemia non-lymphatic haematopathy (including non-anaemic erythrocyte disorders, leukocyte disorders, haemorrhagic disorders and haematological malignancies):**
○ Yes
○ No

### D. Obstetric and pregnancy-related medical conditions

*20 risk scoring indicators*

**D-1. Uterine fibroids:**
○ Yes
○ No

**D-2. Uterine malformation:**
○ Yes
○ No

**D-3. Uterine rupture:**
○ Yes
○ No

**D-4. Congenital uterine anomalies:**
○ Yes
○ No

**D-5. Uterine polyps:**
○ Yes
○ No

**D-6. Cervical incompetence:**
○ Yes
○ No

**D-7. History of cervical surgery:**
○ Yes
○ No

**D-8. Cervical polyps:**

○ Yes

○ No

**D-9. History of cervical cone biopsy:**

○ Yes

○ No

**D-10. Placenta implantation:**

○ Yes

○ No

**D-11. Chorioamnionitis:**

○ Yes

○ No

**D-12. Placental adhesion:**

○ Yes

○ No

**D-13. Placental abruption:**

○ Yes

○ No

**D-14. Tubal serous cystadenoma:**

○ Yes

○ No

**D-15. Ovarian cysts:**

○ Yes

○ No

**D-16. Polycystic ovary syndrome (PCOS):**

○ Yes

○ No

**D-17. Vaginitis:**

○ Yes

○ No

**D-18. Umbilical cord abnormalities:**

○ Yes

○ No

**D-19. Syphilis history:**

○ Yes

○ No

**D-20. Singleton or multiple pregnancies:**

○ Singleton

○ Twins

○ Triplets or more

### E. Other metabolic, immune, and mental conditions

*29 risk scoring indicators*

**E-1. Primary (non-pregnancy-induced) Type-II diabetes (current condition or history):**

○ Yes

○ No

**E-2. Gestational diabetes mellitus (GDM):**

○ Yes

○ No

**E-3. Hepatitis B (current condition or history):**

○ Yes

○ No

**E-4. Hepatitis C (current condition or history):**

○ Yes

○ No

**E-5. Gallstones or cholecystitis (current condition or history):**

○ Yes

○ No

**E-6. Single-symptom intrahepatic cholestasis of pregnancy (ICP-S):**

○ Yes

○ No

**E-7. Multi-symptomatic intrahepatic cholestasis of pregnancy (ICP-M):**

○ Yes

○ No

**E-8. Gallbladder disease (current condition or history):**

○ Yes

○ No

**E-9. History of cholestasis:**
○ Yes
○ No

**E-10. Fatty liver disease (FLD, hepatic steatosis):**
○ Yes
○ No

**E-11. Hepatic haemangioma:**
○ Yes
○ No

**E-12. Hyperthyroidism:**
○ Yes
○ No

**E-13. Hypothyroidism:**
○ Yes
○ No

**E-14. Hashimoto's thyroiditis:**
○ Yes
○ No

**E-15. Thyroid nodules:**
○ Yes
○ No

**E-16. History of thyroid surgery:**
○ Yes
○ No

**E-17. Total thyroidectomy:**
○ Yes
○ No

**E-18. Subtotal thyroidectomy:**
○ Yes
○ No

**E-19. History of thyroid malignancy:**
○ Yes
○ No

**E-20. Group B streptococcus carrier:**

○ Yes

○ No

**E-21. Thrombophilia:**

○ Yes

○ No

**E-22. Antiphospholipid syndrome (APS):**

○ Yes

○ No

**E-23. Systemic lupus erythematosus (SLE):**

○ Yes

○ No

**E-24. Endometriosis:**

○ Yes

○ No

**E-25. Renal diseases:**

○ Yes

○ No

**E-26. Ankylosing spondylitis:**

○ Yes

○ No

**E-27. Epilepsy:**

○ Yes

○ No

**E-28. Psycho-behavioural abnormality:**

○ Yes

○ No

**E-29. Depression:**

○ Yes

○ No

### F. Environmental exposure levels:

*15 risk scoring indicators*

*Note: This section does not require active input from the participants. The cloud computing system is designed to use a geographic information coding system to convert the addresses provided by the participants into latitude and longitude coordinates and then map them onto the ambient environmental tracking database. Subsequently, the cloud computing system will automatically calculate the environmental exposure levels for specific gestational periods and match them with the predefined multiple-choice questionnaire options.*

**F-1.1. $PM_{2.5}$ exposure, 12–6 months before conception:** ___________ μg/m$^3$
○ ≤35.0
○ 35.1–40.0
○ 40.1–45.0
○ >45.0

**F-1.2. $PM_{2.5}$ exposure, 6–3 months before conception:** ___________ μg/m$^3$
○ ≤35.0
○ 35.1–40.0
○ 40.1–45.0
○ >45.0

**F-1.3. $PM_{2.5}$ exposure, 3 months before conception:** ___________ μg/m$^3$
○ ≤35.0
○ 35.1–40.0
○ 40.1–45.0
○ >45.0

**F-1.4. $PM_{2.5}$ exposure, first trimester (1–3 months since conception):** ___________ μg/m$^3$
○ ≤35.0
○ 35.1–40.0
○ 40.1–45.0
○ >45.0

**F-1.5. $PM_{2.5}$ exposure, second trimester (4–6 months since conception):** ___________ μg/m$^3$
○ ≤35.0
○ 35.1–40.0
○ 40.1–45.0
○ >45.0

**F-2.1. $O_3$ exposure, 12–6 months before conception:** ___________ ppb
○ ≤40.0
○ 40.1–45.0
○ 45.1–50.0
○ >50.0

**F-2.2. $O_3$ exposure, 6–3 months before conception:** ___________ ppb
○ ≤40.0

○ 40.1–45.0
○ 45.1–50.0
○ >50.0

**F-2.3. $O_3$ exposure, 3 months before conception:** ___________ ppb
○ ≤40.0
○ 40.1–45.0
○ 45.1–50.0
○ >50.0

**F-2.4. $O_3$ exposure, first trimester (1–3 months since conception):** ___________ ppb
○ ≤40.0
○ 40.1–45.0
○ 45.1–50.0
○ >50.0

**F-2.5. $O_3$ exposure, second trimester (4–6 months since conception):** ___________ ppb
○ ≤40.0
○ 40.1–45.0
○ 45.1–50.0
○ >50.0

**F-3.1. Greenness exposure within 500-m residential area, 12–6 months before conception:**
___________ EVI
○ ≤0.150
○ 0.151–0.200
○ 0.201–0.250
○ >0.250

**F-3.2. Greenness exposure within 500-m residential area, 6–3 months before conception:**
___________ EVI
○ ≤0.150
○ 0.151–0.200
○ 0.201–0.250
○ >0.250

**F-3.3. Greenness exposure within 500-m residential area, 3 months before conception:**
___________ EVI
○ ≤0.150
○ 0.151–0.200
○ 0.201–0.250
○ >0.250

**F-3.4. Greenness exposure within 500-m residential area, first trimester (1–3 months since**

**conception):** ___________ EVI

○ ≤0.150

○ 0.151–0.200

○ 0.201–0.250

○ >0.250

**F-3.5. Greenness exposure within 500-m residential area, second trimester (4–6 months since conception):** ___________ EVI

○ ≤0.150

○ 0.151–0.200

○ 0.201–0.250

○ >0.250

### Risk evaluation summary

**Risk score for obstetric adverse pregnancy outcomes:** ____________

**Risk level for obstetric adverse pregnancy outcomes: Normal/Transitional/Risked**

Socioeconomic risk proportion: ___________ %

Cardiovascular and haematological disease risk proportion: ___________ %

Obstetric and pregnancy-related disease risk proportion: ___________ %

Other disease risk proportion: ___________ %

Environmental risk proportion: ___________ %

**Risk score for neonatal cardiovascular diseases:** ____________

**Risk level for neonatal cardiovascular diseases: Normal/Transitional/Risked**

Socioeconomic risk proportion: ___________ %

Cardiovascular and haematological disease risk proportion: ___________ %

Obstetric and pregnancy-related disease risk proportion: ___________ %

Other disease risk proportion: ___________ %

Environmental risk proportion: ___________ %

**Please take caution that the results of the self-administrated risk-assessment questionnaire are for reference only and cannot be used as a basis for clinical diagnosis. This questionnaire is intended to help pregnant women and their families gain a more comprehensive understanding of their pregnancy risk factors. Being categorised as “Normal” does not guarantee the absence of pregnancy complications, and conversely, being categorised as “Risked” does not guarantee the occurrence of adverse outcomes. Pregnant women and families must adhere to medical advice, and clinical diagnoses should be based on real-time changes in clinical symptoms as determined by healthcare professionals.**

**Supplementary Table S2 | Scoring for risk factors of self-directed questionnaire for early-stage risk forecasting of obstetric adverse pregnancy outcomes and neonatal congenital cardiovascular diseases.**

Since two categories of adverse health outcomes are independently predicted, each risk factor is assigned two risk scores of weighting: one for obstetric adverse pregnancy outcomes (*marked in Blue on the left*) and the other for neonatal congenital cardiovascular diseases (*marked in Grey on the right*).

### A. Residence information for the current pregnancy

*Section A is not directly involved for risk scoring, thus omitted here*

### B. Basic socioeconomic characteristics

*10 risk scoring features*

**B-1. Age at the current pregnancy:**

○ ≤29
○ 29–39 2.4 | 0.8
○ >39 3.2 | 1.4

**B-2. Hukou category (household registration type):**

○ Rural 1.6 | 0.9
○ Rural-to-urban migration 0.8 | 0.4
○ Urban

**B-3. Highest educational attainment level:**

○ Elementary school or below 0.7 | 0.9
○ High school or technical school 0.5 | 0.6
○ Undergraduate or college 0.3 | 0.3
○ Postgraduate or higher

**B-4. Household income level:**

○ ≤5,000 0.5 | 2.4
○ 5,000–20,000 0.3 | 1.6
○ 20,000–50,000 0.2 | 0.8
○ >50,000

**B-5. Smoking habit:**

○ No
○ Passive smoking (current or history) 3.3 | 0.8
○ Active smoking (current or history) 4.6 | 1.4

**B-6. Alcohol consumption habit:**

○ No
○ Yes (current or history) 1.6 | 0.8

**B-10. Pre-pregnancy BMI (automatically calculated):**

○ ≤18.5 0.6 | 0.5

○ 18.5–23.9

○ >23.9 0.3 | 0.2

**B-11. BMI upon delivery (automatically calculated):**

○ ≤29.8 2.1 | 1.0

○ 29.8–32.8

○ >32.8 1.2 | 0.5

**B-12. Parity of the current pregnancy:**

○ 1

○ 2–3 0.1 | 0.5

○ >3 0.2 | 0.9

**B-13. Gravidity of the current pregnancy:**

○ 1

○ 2–3 0.2 | 0.1

○ >3 0.4 | 0.1

### C. Cardiovascular and haematological conditions diagnosed at the current pregnancy

*11 risk scoring indicators*

**C-1. Nutritional anaemia:**

○ Yes 3.2 | 2.1

○ No

**C-2. Haemolytic anaemia:**

○ Yes 1.2 | 2.6

○ No

**C-3. Congenital heart disease (CHD):**

○ Yes 5.8 | 5.6

○ No

**C-4. Pulmonary arterial hypertension (PAH):**

○ Yes 5.4 | 1.6

○ No

**C-5. Primary non-pregnancy-induced hypertension:**

○ Yes 0.5 | 2.1

○ No

**C-6. Pregnancy-induced hypertension:**

○ Yes 10.0 | 1.6

○ No

**C-7. Preeclampsia:**

○ Yes 40.0 | 12.7

○ No

**C-8. Cardiac insufficiency:**

○ Yes 10.1 | 4.9

○ No

**C-9. Arrhythmia:**

○ Yes 3.2 | 1.4

○ No

**C-10. Lymphatic system diseases:**

○ Yes 6.4 | 1.2

○ No

**C-11. Non-anaemia non-lymphatic haematopathy (including non-anaemic erythrocyte disorders, leukocyte disorders, haemorrhagic disorders and haematological malignancies):**

○ Yes 0.6 | 1.9

○ No

### D. Obstetric and pregnancy-related medical conditions

*20 risk scoring indicators*

**D-1. Uterine fibroids:**

○ Yes 0.1 | 0.1

○ No

**D-2. Uterine malformation:**

○ Yes 0.1 | 0.1

○ No

**D-3. Uterine rupture:**

○ Yes 0.3 | 0.1

○ No

**D-4. Congenital uterine anomalies:**

○ Yes 0.3 | 0.6

○ No

**D-5. Uterine polyps:**

○ Yes 0.1 | 0.1

○ No

**D-6. Cervical incompetence:**

○ Yes 1.0 | 0.8

○ No

**D-7. History of cervical surgery:**

○ Yes 0.2 | 0.6

○ No

**D-8. Cervical polyps:**

○ Yes 0.3 | 0.6

○ No

**D-9. History of cervical conisation (LEEP):**

○ Yes 0.3 | 0.6

○ No

**D-10. Placenta accreta, increta, or percreta:**

○ Yes 1.9 | 1.4

○ No

**D-11. Chorioamnionitis and endometritis:**

○ Yes 0.2 | 0.4

○ No

**D-12. Placental adhesion:**

○ Yes 0.1 | 0.5

○ No

**D-13. Placental abruption:**

○ Yes 0.8 | 1.6

○ No

**D-14. Mesosalpinx cyst:**

○ Yes 0.1 | 0.1

○ No

**D-15. Ovarian cysts:**

○ Yes 0.1 | 0.1

○ No

**D-16. Polycystic ovary syndrome (PCOS):**

○ Yes 0.1 | 0.3

○ No

**D-17. Vaginitis:**

○ Yes 1.8 | 0.1

○ No

**D-18. Umbilical cord abnormalities:**

○ Yes 0.1 | 0.1

○ No

**D-19. History of Syphilis:**

○ Yes 0.1 | 0.4

○ No

**D-20. Singleton or multiple pregnancies:**

○ Singleton

○ Twins 30.0 | 0.1

○ Triplets or more 30.0 | 0.2

### E. Other metabolic, immune, and mental conditions

*29 risk scoring indicators*

**E-1. Primary (non-pregnancy-induced) Type-II diabetes (current condition or history):**

○ Yes 0.1 | 0.3

○ No

**E-2. Gestational diabetes mellitus (GDM):**

○ Yes 1.2 | 1.9

○ No

**E-3. Hepatitis B (current condition or history):**

○ Yes 0.1 | 0.1

○ No

**E-4. Hepatitis C or E (current condition or history):**

○ Yes 0.1 | 0.3

○ No

**E-5. Gallstones or cholecystitis (current condition or history):**

○ Yes 0.1 | 0.4

○ No

**E-6. Single-symptom intrahepatic cholestasis of pregnancy (ICP-S):**

○ Yes 0.1 | 0.1

○ No

**E-7. Multi-symptomatic intrahepatic cholestasis of pregnancy (ICP-M):**

○ Yes 1.2 | 0.4

○ No

**E-8. Gallbladder disease (current condition or history):**

○ Yes 0.1 | 0.4

○ No

**E-9. Primary or history of cholestasis:**

○ Yes 0.5 | 0.5

○ No

**E-10. Fatty liver disease (FLD, hepatic steatosis):**

○ Yes 0.4 | 0.3

○ No

**E-11. Hepatic haemangioma:**

○ Yes 0.1 | 0.5

○ No

**E-12. Hyperthyroidism:**

○ Yes 0.1 | 0.1

○ No

**E-13. Hypothyroidism:**

○ Yes 0.1 | 0.1

○ No

**E-14. Hashimoto's thyroiditis:**

○ Yes 0.1 | 0.1

○ No

**E-15. Thyroid nodules:**

○ Yes 0.2 | 0.4

○ No

**E-16. History of thyroid surgery:**

○ Yes 0.1 | 0.3

○ No

**E-17. Total thyroidectomy:**

○ Yes 0.3 | 1.2

○ No

**E-18. Subtotal thyroidectomy:**

○ Yes 0.1 | 0.5

○ No

**E-19. History of thyroid malignancy:**

○ Yes 0.1 | 0.1

○ No

**E-20. Group B streptococcus carrier:**

○ Yes 0.1 | 0.1

○ No

**E-21. Thrombophilia:**

○ Yes 0.1 | 0.1

○ No

**E-22. Antiphospholipid syndrome (APS):**

○ Yes 0.1 | 0.1

○ No

**E-23. Systemic lupus erythematosus (SLE):**

○ Yes 0.4 | 0.7

○ No

**E-24. Endometriosis:**

○ Yes 0.1 | 0.1

○ No

**E-25. Renal diseases:**

○ Yes 0.1 | 0.1

○ No

**E-26. Ankylosing spondylitis:**

○ Yes 0.1 | 0.1

○ No

**E-27. Epilepsy:**

○ Yes 0.1 | 0.3

○ No

**E-28. Psycho-behavioural abnormality:**
○ Yes 0.1 | 0.1
○ No

**E-29. Depression:**
○ Yes 0.3 | 0.7
○ No

### F. Environmental exposure levels:

*15 risk scoring indicators*

**F-1.1. $PM_{2.5}$ exposure, 12–6 months before conception:** ___________ μg/m$^3$
○ ≤35.0
○ 35.1–40.0 0.2 | 1.0
○ 40.1–45.0 0.4 | 1.8
○ >45.0 0.7 | 2.6

**F-1.2. $PM_{2.5}$ exposure, 6–3 months before conception:** ___________ μg/m$^3$
○ ≤35.0
○ 35.1–40.0 1.0 | 0.6
○ 40.1–45.0 1.8 | 1.0
○ >45.0 2.7 | 1.6

**F-1.3. $PM_{2.5}$ exposure, 3 months before conception:** ___________ μg/m$^3$
○ ≤35.0
○ 35.1–40.0 0.6 | 3.0
○ 40.1–45.0 1.0 | 5.9
○ >45.0 1.6 | 8.8

**F-1.4. $PM_{2.5}$ exposure, first trimester (1–3 months since conception):** ___________ μg/m$^3$
○ ≤35.0
○ 35.1–40.0 2.6 | 1.0
○ 40.1–45.0 5.5 | 2.0
○ >45.0 7.9 | 2.8

**F-1.5. $PM_{2.5}$ exposure, second trimester (4–6 months since conception):** ___________ μg/m$^3$
○ ≤35.0
○ 35.1–40.0 0.9 | 0.4
○ 40.1–45.0 1.9 | 0.8
○ >45.0 2.8 | 1.2

**F-2.1. $O_3$ exposure, 12–6 months before conception:** ___________ ppb

○ ≤40.0
○ 40.1–45.0 0.6 | 0.1
○ 45.1–50.0 1.2 | 0.1
○ >50.0 1.8 | 0.1

**F-2.2. $O_3$ exposure, 6–3 months before conception:** ___________ ppb

○ ≤40.0
○ 40.1–45.0 1.5 | 1.0
○ 45.1–50.0 3.2 | 2.1
○ >50.0 4.6 | 3.1

**F-2.3. $O_3$ exposure, 3 months before conception:** ___________ ppb

○ ≤40.0
○ 40.1–45.0 0.5 | 0.4
○ 45.1–50.0 1.0 | 0.8
○ >50.0 1.5 | 1.0

**F-2.4. $O_3$ exposure, first trimester (1–3 months since conception):** ___________ ppb

○ ≤40.0
○ 40.1–45.0 0.8 | 1.0
○ 45.1–50.0 1.5 | 2.2
○ >50.0 2.2 | 3.0

**F-2.5. $O_3$ exposure, second trimester (4–6 months since conception):** ___________ ppb

○ ≤40.0
○ 40.1–45.0 0.6 | 0.6
○ 45.1–50.0 1.2 | 1.1
○ >50.0 1.7 | 1.6

**F-3.1. Greenness exposure within 500-m residential area, 12–6 months before conception:** ___________ EVI

○ ≤0.150 0.5 | 0.6
○ 0.151–0.200 0.3 | 0.4
○ 0.201–0.250 0.1 | 0.2
○ >0.250

**F-3.2. Greenness exposure within 500-m residential area, 6–3 months before conception:** ___________ EVI

○ ≤0.150 2.9 | 2.0
○ 0.151–0.200 2.0 | 1.4
○ 0.201–0.250 1.0 | 0.8
○ >0.250

**F-3.3. Greenness exposure within 500-m residential area, 3 months before conception:**

___________ EVI

○ ≤0.150 0.8 | 1.2

○ 0.151–0.200 0.5 | 0.8

○ 0.201–0.250 0.3 | 0.4

○ >0.250

**F-3.4. Greenness exposure within 500-m residential area, first trimester (1–3 months since conception):** ___________ EVI

○ ≤0.150 3.2 | 1.1

○ 0.151–0.200 2.2 | 0.8

○ 0.201–0.250 1.2 | 0.4

○ >0.250

**F-3.5. Greenness exposure within 500-m residential area, second trimester (4–6 months since conception):** ___________ EVI

○ ≤0.150 2.8 | 1.9

○ 0.151–0.200 1.8 | 1.3

○ 0.201–0.250 1.0 | 0.6

○ >0.250

### Risk evaluation summary and categorisation

For obstetric adverse pregnancy outcomes, the algorithm specifies that a total risk score greater than 58.4 falls into the "Risked" category, suggesting that pregnant individuals may require special medical monitoring. Risk scores greater than 41.2 are placed in the "Transitional" category, indicating a higher risk compared to the average population. Scores below 41.2 are categorised as "Normal", suggesting that pregnant individuals have avoided most risk factors. For neonatal cardiovascular diseases, the algorithm specifies that a total risk score greater than 56.7 falls into the "Risked" category; risk scores greater than 45.0 are classified as "Transitional"; scores below 45.0 are identified as "Normal". These groupings are entirely based on statistical inference, and thus, the results are for reference purposes only.

**Supplementary Table S3 | A sample filled self-directed questionnaire with the highest risk score of obstetric adverse pregnancy outcomes.**

### A. Residence information for the current pregnancy

*Section A is not directly involved for risk scoring, thus omitted here*

Internal ZEBRA series No.: 00000648

### B. Basic socioeconomic characteristics

*10 risk scoring features*

**B-1. Age at the current pregnancy:**

○ ≤29

√ 29–39 **2.4**

○ >39

**B-2. Hukou category (household registration type):**

○ Rural

√ Rural-to-urban migration **0.8**

○ Urban

**B-3. Highest educational attainment level:**

√ Elementary school or below **0.7**

○ High school or technical school

○ Undergraduate or college

○ Postgraduate or higher

**B-4. Household income level:**

○ ≤5,000

√ 5,000–20,000 **0.3**

○ 20,000–50,000

○ >50,000

**B-5. Smoking habit:**

○ No

√ Passive smoking (current or history) **3.3**

○ Active smoking (current or history)

**B-6. Alcohol consumption habit:**

√ No

○ Yes (current or history)

**B-10. Pre-pregnancy BMI (automatically calculated):**

○ ≤18.5

○ 18.5–23.9

√ >23.9 0.3

**B-11. BMI upon delivery (automatically calculated):**

○ ≤29.8

○ 29.8–32.8

√ >32.8 1.2

**B-12. Parity of the current pregnancy: 3**

○ 1

√ 2–3 0.1

○ >3

**B-13. Gravidity of the current pregnancy: 6**

○ 1

○ 2–3

√ >3 0.4

### C. Cardiovascular and haematological conditions diagnosed at the current pregnancy

*11 risk scoring indicators*

**C-1. Nutritional anaemia:**

○ Yes

√ No

**C-2. Haemolytic anaemia:**

○ Yes

√ No

**C-3. Congenital heart disease (CHD):**

○ Yes

√ No

**C-4. Pulmonary arterial hypertension (PAH):**

√ Yes 5.4

○ No

**C-5. Primary non-pregnancy-induced hypertension:**

√ Yes 0.5

○ No

**C-6. Pregnancy-induced hypertension:**

√ Yes 10.0

○ No

**C-7. Preeclampsia:**

√ Yes 40.0

○ No

**C-8. Cardiac insufficiency:**

○ Yes

√ No

**C-9. Arrhythmia:**

○ Yes

√ No

**C-10. Lymphatic system diseases:**

○ Yes

√ No

**C-11. Non-anaemia non-lymphatic haematopathy (including non-anaemic erythrocyte disorders, leukocyte disorders, haemorrhagic disorders and haematological malignancies):**

○ Yes

√ No

### D. Obstetric and pregnancy-related medical conditions

*20 risk scoring indicators*

**D-1. Uterine fibroids:**

○ Yes

√ No

**D-2. Uterine malformation:**

○ Yes

√ No

**D-3. Uterine rupture:**

○ Yes

√ No

**D-4. Congenital uterine anomalies:**

○ Yes

√ No

**D-5. Uterine polyps:**

○ Yes

√ No

**D-6. Cervical incompetence:**

○ Yes

√ No

**D-7. History of cervical surgery:**

○ Yes

√ No

**D-8. Cervical polyps:**

○ Yes

√ No

**D-9. History of cervical conisation (LEEP):**

○ Yes

√ No

**D-10. Placenta accreta, increta, or percreta:**

○ Yes

√ No

**D-11. Chorioamnionitis and endometritis:**

○ Yes

√ No

**D-12. Placental adhesion:**

○ Yes

√ No

**D-13. Placental abruption:**

○ Yes

√ No

**D-14. Mesosalpinx cyst:**

○ Yes

○ No

**D-15. Ovarian cysts:**

○ Yes

√ No

**D-16. Polycystic ovary syndrome (PCOS):**

○ Yes

√ No

**D-17. Vaginitis:**

○ Yes

√ No

**D-18. Umbilical cord abnormalities:**

○ Yes

√ No

**D-19. History of Syphilis:**

○ Yes

√ No

**D-20. Singleton or multiple pregnancies:**

√ Singleton

○ Twins

○ Triplets or more

### E. Other metabolic, immune, and mental conditions

*29 risk scoring indicators*

**E-1. Primary (non-pregnancy-induced) Type-II diabetes (current condition or history):**

○ Yes

√ No

**E-2. Gestational diabetes mellitus (GDM):**

○ Yes

√ No

**E-3. Hepatitis B (current condition or history):**

○ Yes

√ No

**E-4. Hepatitis C or E (current condition or history):**

○ Yes

√ No

**E-5. Gallstones or cholecystitis (current condition or history):**

○ Yes

√ No

**E-6. Single-symptom intrahepatic cholestasis of pregnancy (ICP-S):**

○ Yes
√ No

**E-7. Multi-symptomatic intrahepatic cholestasis of pregnancy (ICP-M):**
√ Yes 1.2
○ No

**E-8. Gallbladder disease (current condition or history):**
○ Yes
√ No

**E-9. Primary or history of cholestasis:**
√ Yes 0.5
○ No

**E-10. Fatty liver disease (FLD, hepatic steatosis):**
○ Yes
√ No

**E-11. Hepatic haemangioma:**
○ Yes
√ No

**E-12. Hyperthyroidism:**
○ Yes
√ No

**E-13. Hypothyroidism:**
○ Yes
√ No

**E-14. Hashimoto's thyroiditis:**
○ Yes
√ No

**E-15. Thyroid nodules:**
○ Yes
√ No

**E-16. History of thyroid surgery:**
○ Yes
√ No

**E-17. Total thyroidectomy:**

○ Yes
√ No

**E-18. Subtotal thyroidectomy:**
○ Yes
√ No

**E-19. History of thyroid malignancy:**
○ Yes
√ No

**E-20. Group B streptococcus carrier:**
○ Yes
√ No

**E-21. Thrombophilia:**
√ Yes 0.1
○ No

**E-22. Antiphospholipid syndrome (APS):**
○ Yes
√ No

**E-23. Systemic lupus erythematosus (SLE):**
○ Yes
√ No

**E-24. Endometriosis:**
√ Yes 0.1
○ No

**E-25. Renal diseases:**
○ Yes
√ No

**E-26. Ankylosing spondylitis:**
○ Yes
√ No

**E-27. Epilepsy:**
○ Yes
√ No

**E-28. Psycho-behavioural abnormality:**

√ Yes 0.1

○ No

**E-29. Depression:**

○ Yes

√ No

### F. Environmental exposure levels:

*15 risk scoring indicators*

**F-1.1. $PM_{2.5}$ exposure, 12–6 months before conception:** 48.2 μg/m³

○ ≤35.0

○ 35.1–40.0

○ 40.1–45.0

√ >45.0 0.7

**F-1.2. $PM_{2.5}$ exposure, 6–3 months before conception:** 35.9 μg/m³

○ ≤35.0

√ 35.1–40.0 1.0

○ 40.1–45.0

○ >45.0

**F-1.3. $PM_{2.5}$ exposure, 3 months before conception:** 29.2 μg/m³

√ ≤35.0

○ 35.1–40.0

○ 40.1–45.0

○ >45.0

**F-1.4. $PM_{2.5}$ exposure, first trimester (1–3 months since conception):** 52.0 μg/m³

○ ≤35.0

○ 35.1–40.0

○ 40.1–45.0

√ >45.0 7.0

**F-1.5. $PM_{2.5}$ exposure, second trimester (4–6 months since conception):** 63.1 μg/m³

○ ≤35.0

○ 35.1–40.0

○ 40.1–45.0

√ >45.0 2.8

**F-2.1. $O_3$ exposure, 12–6 months before conception:** 49.6 ppb

○ ≤40.0

○ 40.1–45.0

√ 45.1–50.0 **1.2**
○ >50.0

**F-2.2. $O_3$ exposure, 6–3 months before conception:** __68.1__ ppb
○ ≤40.0
○ 40.1–45.0
○ 45.1–50.0
√ >50.0 **4.6**

**F-2.3. $O_3$ exposure, 3 months before conception:** __66.3__ ppb
○ ≤40.0
○ 40.1–45.0
○ 45.1–50.0
√ >50.0 **1.5**

**F-2.4. $O_3$ exposure, first trimester (1–3 months since conception):** __33.3__ ppb
√ ≤40.0
○ 40.1–45.0
○ 45.1–50.0
○ >50.0

**F-2.5. $O_3$ exposure, second trimester (4–6 months since conception):** __31.2__ ppb
√ ≤40.0
○ 40.1–45.0
○ 45.1–50.0
○ >50.0

**F-3.1. Greenness exposure within 500-m residential area, 12–6 months before conception:** __0.143__ EVI
√ ≤0.150 **0.5**
○ 0.151–0.200
○ 0.201–0.250
○ >0.250

**F-3.2. Greenness exposure within 500-m residential area, 6–3 months before conception:** __0.156__ EVI
○ ≤0.150
√ 0.151–0.200 **2.0**
○ 0.201–0.250
○ >0.250

**F-3.3. Greenness exposure within 500-m residential area, 3 months before conception:** __0.164__ EVI
○ ≤0.150

√ 0.151–0.200 0.5
○ 0.201–0.250
○ >0.250

**F-3.4. Greenness exposure within 500-m residential area, first trimester (1–3 months since conception):** 0.137 EVI
√ ≤0.150 3.2
○ 0.151–0.200
○ 0.201–0.250
○ >0.250

**F-3.5. Greenness exposure within 500-m residential area, second trimester (4–6 months since conception):** 0.092 EVI
√ ≤0.150 2.8
○ 0.151–0.200
○ 0.201–0.250
○ >0.250

**Clinical diagnosis: spontaneous preterm birth and low birth weight neonate.**

### Risk evaluation summary

**Risk score for obstetric adverse pregnancy outcomes:** 96.1
**Risk level for obstetric adverse pregnancy outcomes: Risked**
Socioeconomic risk proportion: 9.5/16.1 = 59.0 %
Cardiovascular and haematological disease risk proportion: 55.9/86.4 = 64.7 %
Obstetric and pregnancy-related disease risk proportion: 0/40.7 = 0 %
Other disease risk proportion: 2.0/6.6 = 30.0 %
Environmental risk proportion: 28.7/37.7 = 76.1 %

**Supplementary Table S4 | A sample filled self-directed questionnaire with the highest risk score of neonatal cardiovascular diseases.**

### A. Residence information for the current pregnancy

*Section A is not directly involved for risk scoring, thus omitted here*

Internal ZEBRA series No.: 00120789

### B. Basic socioeconomic characteristics

*10 risk scoring features*

**B-1. Age at the current pregnancy:**

○ ≤29

√ 29–39 0.8

○ >39

**B-2. Hukou category (household registration type):**

√ Rural 0.9

○ Rural-to-urban migration

○ Urban

**B-3. Highest educational attainment level:**

○ Elementary school or below

√ High school or technical school 0.6

○ Undergraduate or college

○ Postgraduate or higher

**B-4. Household income level:**

○ ≤5,000

√ 5,000–20,000 1.6

○ 20,000–50,000

○ >50,000

**B-5. Smoking habit:**

√ No

○ Passive smoking (current or history)

○ Active smoking (current or history)

**B-6. Alcohol consumption habit:**

√ No

○ Yes (current or history)

**B-10. Pre-pregnancy BMI (automatically calculated):**

○ ≤18.5

○ 18.5–23.9

√ >23.9 0.2

**B-11. BMI upon delivery (automatically calculated):**

○ ≤29.8

○ 29.8–32.8

√ >32.8 0.5

**B-12. Parity of the current pregnancy:**

√ 1

○ 2–3

○ >3

**B-13. Gravidity of the current pregnancy:**

○ 1

√ 2–3 0.1

○ >3

### C. Cardiovascular and haematological conditions diagnosed at the current pregnancy

*11 risk scoring indicators*

**C-1. Nutritional anaemia:**

○ Yes

√ No

**C-2. Haemolytic anaemia:**

○ Yes

√ No

**C-3. Congenital heart disease (CHD):**

○ Yes

√ No

**C-4. Pulmonary arterial hypertension (PAH):**

○ Yes

√ No

**C-5. Primary non-pregnancy-induced hypertension:**

√ Yes 2.1

○ No

**C-6. Pregnancy-induced hypertension:**

√ Yes 1.6

○ No

**C-7. Preeclampsia:**

√ Yes 12.7

○ No

**C-8. Cardiac insufficiency:**

√ Yes 4.9

○ No

**C-9. Arrhythmia:**

√ Yes 1.4

○ No

**C-10. Lymphatic system diseases:**

○ Yes

√ No

**C-11. Non-anaemia non-lymphatic haematopathy (including non-anaemic erythrocyte disorders, leukocyte disorders, haemorrhagic disorders and haematological malignancies):**

√ Yes 1.9

○ No

### D. Obstetric and pregnancy-related medical conditions

*20 risk scoring indicators*

**D-1. Uterine fibroids:**

○ Yes

√ No

**D-2. Uterine malformation:**

○ Yes

√ No

**D-3. Uterine rupture:**

○ Yes

√ No

**D-4. Congenital uterine anomalies:**

○ Yes

√ No

**D-5. Uterine polyps:**

○ Yes

√ No

**D-6. Cervical incompetence:**

○ Yes

√ No

**D-7. History of cervical surgery:**

√ Yes 0.6

○ No

**D-8. Cervical polyps:**

√ Yes 0.6

○ No

**D-9. History of cervical conisation (LEEP):**

○ Yes

√ No

**D-10. Placenta accreta, increta, or percreta:**

√ Yes 1.4

○ No

**D-11. Chorioamnionitis and endometritis:**

○ Yes

√ No

**D-12. Placental adhesion:**

○ Yes

√ No

**D-13. Placental abruption:**

√ Yes 1.6

○ No

**D-14. Mesosalpinx cyst:**

○ Yes

√ No

**D-15. Ovarian cysts:**

○ Yes

√ No

**D-16. Polycystic ovary syndrome (PCOS):**

○ Yes

√ No

**D-17. Vaginitis:**

○ Yes

√ No

**D-18. Umbilical cord abnormalities:**

○ Yes

√ No

**D-19. History of Syphilis:**

○ Yes

√ No

**D-20. Singleton or multiple pregnancies:**

√ Singleton

○ Twins

○ Triplets or more

### E. Other metabolic, immune, and mental conditions

*29 risk scoring indicators*

**E-1. Primary (non-pregnancy-induced) Type-II diabetes (current condition or history):**

○ Yes

√ No

**E-2. Gestational diabetes mellitus (GDM):**

○ Yes

√ No

**E-3. Hepatitis B (current condition or history):**

○ Yes

√ No

**E-4. Hepatitis C or E (current condition or history):**

○ Yes

√ No

**E-5. Gallstones or cholecystitis (current condition or history):**

○ Yes

√ No

**E-6. Single-symptom intrahepatic cholestasis of pregnancy (ICP-S):**

○ Yes
√ No

**E-7. Multi-symptomatic intrahepatic cholestasis of pregnancy (ICP-M):**
○ Yes
√ No

**E-8. Gallbladder disease (current condition or history):**
○ Yes
√ No

**E-9. Primary or history of cholestasis:**
○ Yes
√ No

**E-10. Fatty liver disease (FLD, hepatic steatosis):**
○ Yes
√ No

**E-11. Hepatic haemangioma:**
○ Yes
√ No

**E-12. Hyperthyroidism:**
√ Yes 0.1
○ No

**E-13. Hypothyroidism:**
○ Yes
√ No

**E-14. Hashimoto's thyroiditis:**
○ Yes
√ No

**E-15. Thyroid nodules:**
○ Yes
√ No

**E-16. History of thyroid surgery:**
√ Yes 0.3
○ No

**E-17. Total thyroidectomy:**

√ Yes 1.2
○ No

**E-18. Subtotal thyroidectomy:**
○ Yes
√ No

**E-19. History of thyroid malignancy:**
○ Yes
√ No

**E-20. Group B streptococcus carrier:**
○ Yes
√ No

**E-21. Thrombophilia:**
○ Yes
√ No

**E-22. Antiphospholipid syndrome (APS):**
○ Yes
√ No

**E-23. Systemic lupus erythematosus (SLE):**
○ Yes
√ No

**E-24. Endometriosis:**
○ Yes
√ No

**E-25. Renal diseases:**
○ Yes
√ No

**E-26. Ankylosing spondylitis:**
○ Yes
√ No

**E-27. Epilepsy:**
○ Yes
√ No

**E-28. Psycho-behavioural abnormality:**

○ Yes
√ No

**E-29. Depression:**
○ Yes
√ No

### F. Environmental exposure levels:

*15 risk scoring indicators*

**F-1.1. $PM_{2.5}$ exposure, 12–6 months before conception:** __61.0__ μg/m$^3$
○ ≤35.0
○ 35.1–40.0
○ 40.1–45.0
√ >45.0 2.6

**F-1.2. $PM_{2.5}$ exposure, 6–3 months before conception:** __60.3__ μg/m$^3$
○ ≤35.0
○ 35.1–40.0
○ 40.1–45.0
√ >45.0 1.6

**F-1.3. $PM_{2.5}$ exposure, 3 months before conception:** __77.1__ μg/m$^3$
○ ≤35.0
○ 35.1–40.0
○ 40.1–45.0
√ >45.0 8.8

**F-1.4. $PM_{2.5}$ exposure, first trimester (1–3 months since conception):** __73.9__ μg/m$^3$
○ ≤35.0
○ 35.1–40.0
○ 40.1–45.0
√ >45.0 2.8

**F-1.5. $PM_{2.5}$ exposure, second trimester (4–6 months since conception):** __45.9__ μg/m$^3$
○ ≤35.0
○ 35.1–40.0
○ 40.1–45.0
√ >45.0 1.2

**F-2.1. $O_3$ exposure, 12–6 months before conception:** __49.6__ ppb
○ ≤40.0
○ 40.1–45.0

√ 45.1–50.0 0.1

○ >50.0

**F-2.2. $O_3$ exposure, 6–3 months before conception:** 44.2 ppb

○ ≤40.0

√ 40.1–45.0 1.0

○ 45.1–50.0

○ >50.0

**F-2.3. $O_3$ exposure, 3 months before conception:** 27.7 ppb

√ ≤40.0

○ 40.1–45.0

○ 45.1–50.0

○ >50.0

**F-2.4. $O_3$ exposure, first trimester (1–3 months since conception):** 53.2 ppb

○ ≤40.0

○ 40.1–45.0

○ 45.1–50.0

√ >50.0 3.0

**F-2.5. $O_3$ exposure, second trimester (4–6 months since conception):** 82.6 ppb

○ ≤40.0

○ 40.1–45.0

○ 45.1–50.0

√ >50.0 1.6

**F-3.1. Greenness exposure within 500-m residential area, 12–6 months before conception:** 0.209 EVI

○ ≤0.150

○ 0.151–0.200

√ 0.201–0.250 0.2

○ >0.250

**F-3.2. Greenness exposure within 500-m residential area, 6–3 months before conception:** 0.256 EVI

○ ≤0.150

○ 0.151–0.200

○ 0.201–0.250

√ >0.250

**F-3.3. Greenness exposure within 500-m residential area, 3 months before conception:** 0.154 EVI

○ ≤0.150

√ 0.151–0.200 0.8
○ 0.201–0.250
○ >0.250

**F-3.4. Greenness exposure within 500-m residential area, first trimester (1–3 months since conception):** 0.113 EVI

√ ≤0.150 1.1
○ 0.151–0.200
○ 0.201–0.250
○ >0.250

**F-3.5. Greenness exposure within 500-m residential area, second trimester (4–6 months since conception):** 0.198 EVI

○ ≤0.150
√ 0.151–0.200 1.3
○ 0.201–0.250
○ >0.250

**Clinical diagnosis: medically indicated preterm birth, extremely low birth weight neonate, neonatal respiratory distress syndrome, and neonatal cardiac disorders.**

### Risk evaluation summary

**Risk score for obstetric adverse pregnancy outcomes:** 82.5
**Risk level for obstetric adverse pregnancy outcomes: Risked**

Socioeconomic risk proportion: 4.7/10.5 = 44.8 %
Cardiovascular and haematological disease risk proportion: 24.6/37.7 = 65.3 %
Obstetric and pregnancy-related disease risk proportion: 4.2/8.8 = 47.7 %
Other disease risk proportion: 1.6/10.4 = 15.4 %
Environmental risk proportion: 26.1/32.6 = 80.1 %

**Supplementary Table S5 | Sensitivity analysis: Fully adjusted risk associations between environmental exposure and adverse health outcomes by bootstrap resampling.**

A total of five bootstrap subgroups (50% sampling size) are randomly extracted for risk association quantification between adverse reproductive health outcomes, including the maternal pregnancy-induced cardiovascular diseases (PI-CVDs) and adverse pregnancy outcomes (APOs), and three major environmental exposures, as $PM_{2.5}$, $O_3$, and greenness. Relative risks quantified by hazard ratios (HRs) are estimated by fully adjusted Cox regression models. Coefficients of variation in percentage for the log-transformed HRs are calculated to represent the cross-subgroup divergences.

| Environmental exposures | | Maternal PI-CVDs | APOs |
|---|---|---|---|
| **$PM_{2.5}$** (10-μg/$m^3$) | Bootstrap subgroup 1 | 1.069 (1.057–1.082) | 1.017 (1.012–1.022) |
| | Bootstrap subgroup 2 | 1.069 (1.056–1.082) | 1.021 (1.016–1.026) |
| | Bootstrap subgroup 3 | 1.084 (1.071–1.097) | 1.017 (1.012–1.022) |
| | Bootstrap subgroup 4 | 1.070 (1.058–1.083) | 1.014 (1.010–1.019) |
| | Bootstrap subgroup 5 | 1.069 (1.057–1.082) | 1.016 (1.012–1.021) |
| | Coefficient of variation (%) | 7.65% | 12.1% |
| **$O_3$** (10-ppb) | Bootstrap subgroup 1 | 1.030 (1.023–1.038) | 1.018 (1.015–1.021) |
| | Bootstrap subgroup 2 | 1.027 (1.020–1.035) | 1.018 (1.015–1.021) |
| | Bootstrap subgroup 3 | 1.025 (1.018–1.033) | 1.018 (1.015–1.021) |
| | Bootstrap subgroup 4 | 1.031 (1.023–1.038) | 1.017 (1.014–1.020) |
| | Bootstrap subgroup 5 | 1.031 (1.023–1.038) | 1.017 (1.014–1.020) |
| | Coefficient of variation (%) | 7.41% | 3.35% |
| **Greenness** (0.1-EVI) | Bootstrap subgroup 1 | 0.963 (0.939–0.987) | 0.969 (0.960–0.979) |
| | Bootstrap subgroup 2 | 0.967 (0.943–0.991) | 0.967 (0.958–0.976) |
| | Bootstrap subgroup 3 | 0.963 (0.939–0.988) | 0.964 (0.955–0.973) |
| | Bootstrap subgroup 4 | 0.968 (0.944–0.992) | 0.967 (0.958–0.976) |
| | Bootstrap subgroup 5 | 0.962 (0.938–0.986) | 0.963 (0.954–0.972) |
| | Coefficient of variation (%) | 6.82% | 7.23% |

**Supplementary Table S6 | Sensitivity analysis: Fully adjusted risk associations between maternal cardiovascular diseases (primary and pregnancy-induced) and adverse pregnancy outcomes (obstetric and neonatal diseases) by bootstrap resampling.**

A total of five bootstrap subgroups (50% sampling size) are randomly extracted for risk association quantification between adverse pregnancy outcomes, including the obstetric adverse pregnancy outcomes (OAPOs) and neonatal cardiovascular diseases (NCVDs), and three major types of maternal diseases, as PI-CVDs, primary hypertension and primary anaemia (corresponding to Fig. 2). Relative risks quantified by hazard ratios (HRs) are estimated by fully adjusted Cox regression models. Coefficients of variation in percentage for the log-transformed HRs are calculated to represent the cross-subgroup divergences.

| Maternal conditions | | OAPOs | NCVDs |
|---|---|---|---|
| **Primary hypertension** | Bootstrap subgroup 1 | 4.03 (3.58–4.53) | 2.02 (1.38–2.98) |
| | Bootstrap subgroup 2 | 4.28 (3.77–4.86) | 2.33 (1.57–3.45) |
| | Bootstrap subgroup 3 | 4.56 (4.03–5.16) | 2.48 (1.72–3.60) |
| | Bootstrap subgroup 4 | 4.26 (3.76–4.83) | 2.37 (1.63–3.45) |
| | Bootstrap subgroup 5 | 4.70 (4.15–5.31) | 2.57 (1.81–3.66) |
| | Coefficient of variation (%) | 3.69% | 9.62% |
| **Primary anaemia** | Bootstrap subgroup 1 | 1.44 (1.34–1.55) | 1.39 (1.10–1.75) |
| | Bootstrap subgroup 2 | 1.53 (1.43–1.65) | 1.49 (1.19–1.86) |
| | Bootstrap subgroup 3 | 1.46 (1.35–1.57) | 1.41 (1.11–1.78) |
| | Bootstrap subgroup 4 | 1.47 (1.36–1.58) | 1.41 (1.11–1.78) |
| | Bootstrap subgroup 5 | 1.46 (1.36–1.57) | 1.36 (1.07–1.72) |
| | Coefficient of variation (%) | 5.75% | 8.85% |
| **Pregnancy-induced CVDs** | Bootstrap subgroup 1 | 13.1 (11.2–15.5) | 3.35 (2.06–5.43) |
| | Bootstrap subgroup 2 | 12.6 (10.7–14.9) | 2.88 (1.76–4.73) |
| | Bootstrap subgroup 3 | 12.9 (10.9–15.1) | 3.69 (2.37–5.72) |
| | Bootstrap subgroup 4 | 13.6 (11.5–16.0) | 3.38 (2.16–5.29) |
| | Bootstrap subgroup 5 | 12.4 (10.6–14.6) | 3.68 (2.41–5.64) |
| | Coefficient of variation (%) | 1.24% | 7.38% |

**Supplementary Table S7 | Sensitivity analysis: Risk associations between environmental exposure and adverse health outcomes by different combination of multi-factor adjustment.**

All the other covariates were fully adjusted throughout the sensitivity analysis. Coefficients of variation in percentage for the log-transformed HRs are calculated based on multi-exposure models involving $PM_{2.5}$, $O_3$, and greenness at least. Abbreviations: *green* for greenness, *temp* for all temperature-related exposure metrics.

| Studied exposures | Adjusted exposures | Maternal PI-CVDs | APOs |
|---|---|---|---|
| **$PM_{2.5}$** (10-µg/$m^3$) | Null | 1.054 (1.047–1.061) | 1.014 (1.011–1.017) |
| | $O_3$ | 1.073 (1.065–1.081) | 1.024 (1.021–1.027) |
| | greenness | 1.048 (1.041–1.056) | 1.008 (1.005–1.011) |
| | $O_3$, green | 1.068 (1.060–1.077) | 1.018 (1.015–1.022) |
| | $O_3$, green, temp | 1.073 (1.064–1.082) | 1.019 (1.015–1.022) |
| | $O_3$, green, temp, $NO_2$ | 1.073 (1.064–1.082) | 1.019 (1.015–1.022) |
| | $O_3$, green, temp, $NO_2$, $SO_2$ | 1.072 (1.062–1.081) | 1.017 (1.014–1.021) |
| | $O_3$, green, temp, $NO_2$, $SO_2$, CO | 1.073 (1.064–1.082) | 1.015 (1.012–1.019) |
| | Coefficient of variation (%) | 2.35% | 7.87% |
| **$O_3$** (10-ppb) | Null | 1.008 (1.004–1.013) | 1.011 (1.009–1.013) |
| | $PM_{2.5}$ | 1.010 (1.005–1.014) | 1.017 (1.015–1.019) |
| | greenness | 1.014 (1.009–1.020) | 1.012 (1.010–1.014) |
| | $PM_{2.5}$, green | 1.029 (1.024–1.034) | 1.017 (1.014–1.019) |
| | $PM_{2.5}$, green, temp | 1.028 (1.023–1.034) | 1.016 (1.014–1.018) |
| | $PM_{2.5}$, green, temp, $NO_2$ | 1.027 (1.022–1.033) | 1.017 (1.014–1.019) |
| | $PM_{2.5}$, green, temp, $NO_2$, $SO_2$ | 1.027 (1.022–1.033) | 1.017 (1.014–1.019) |
| | $PM_{2.5}$, green, temp, $NO_2$, $SO_2$, CO | 1.027 (1.022–1.033) | 1.017 (1.015–1.019) |
| | Coefficient of variation (%) | 2.51% | 1.40% |
| **Greenness** (0.1-EVI) | Null | 0.931 (0.916–0.947) | 0.955 (0.949–0.961) |
| | $PM_{2.5}$ | 0.964 (0.948–0.981) | 0.960 (0.953–0.966) |
| | $O_3$ | 0.928 (0.913–0.944) | 0.952 (0.946–0.959) |
| | $PM_{2.5}$, $O_3$ | 0.969 (0.952–0.986) | 0.963 (0.957–0.969) |
| | $PM_{2.5}$, $O_3$, temp | 0.968 (0.951–0.985) | 0.962 (0.956–0.969) |
| | $PM_{2.5}$, $O_3$, temp, $NO_2$ | 0.964 (0.948–0.982) | 0.963 (0.956–0.969) |
| | $PM_{2.5}$, $O_3$, temp, $NO_2$, $SO_2$ | 0.964 (0.947–0.981) | 0.967 (0.960–0.973) |
| | $PM_{2.5}$, $O_3$, temp, $NO_2$, $SO_2$, CO | 0.964 (0.948–0.982) | 0.961 (0.954–0.967) |
| | Coefficient of variation (%) | 6.03% | 5.27% |

### Supplementary Table S8 | Sensitivity analysis: Robustness of estimated risk associations at Zhejiang Provincial scale and China nationwide scale.

During the starting phases of ZEBRA maternity cohort-based research, participant enrolment was biased towards collection from within Zhejiang Province. Despite ZEBRA's principle of including all pregnant women and newborns nationwide within the same medical diagnostic system, as of 2022, nearly 90% of participants are from Zhejiang Province. This introduces a potential limitation in the generalisability of the findings from the present study onto a broader population. To address this, a set of sensitivity analyses was conducted: the currently included population was split into two subsets, consisting of pregnant women from within Zhejiang Province and those from outside Zhejiang Province, for all major epidemiological analyses covered in this study. Meta-analysis (Hunter-Schmidt random-effects estimator) was employed to assess the heterogeneity of estimated HR values between the two subgroups, using Higgins $I^2$ as a statistical indicator, with $\alpha$=0.05 set as the critical level for significance.

| Risk factors | Outcomes | Zhejiang Province | Other Provinces | Heterogeneity |
|---|---|---|---|---|
| **$PM_{2.5}$** (10-μg/m$^3$) | **PI-CVDs** | 1.072 (1.062–1.082) | 1.093 (1.058–1.130) | $I^2$ = 22.3%, $p$ = 0.26 |
| **$O_3$** (10-ppb) | **PI-CVDs** | 1.028 (1.022–1.033) | 1.029 (1.008–1.051) | $I^2$ = 0%, $p$ = 0.88 |
| **Greenness** (0.1-EVI) | **PI-CVDs** | 0.963 (0.946–0.981) | 0.980 (0.919–1.046) | $I^2$ = 0%, $p$ = 0.61 |
| **$PM_{2.5}$** (10-μg/m$^3$) | **APOs** | 1.019 (1.015–1.022) | 1.020 (1.007–1.034) | $I^2$ = 0%, $p$ = 0.80 |
| **$O_3$** (10-ppb) | **APOs** | 1.016 (1.014–1.018) | 1.023 (1.014–1.031) | $I^2$ = 55.7%, $p$ = 0.13 |
| **Greenness** (0.1-EVI) | **APOs** | 0.965 (0.958–0.971) | 0.949 (0.931–0.968) | $I^2$ = 54.6%, $p$ = 0.14 |
| **Prim hypertension** | **OAPOs** | 3.83 (3.52–4.18) | 3.61 (2.60–5.00) | $I^2$ = 0%, $p$ = 0.72 |
| **Prim anaemia** | **OAPOs** | 1.42 (1.34–1.50) | 1.53 (1.24–1.88) | $I^2$ = 0%, $p$ = 0.51 |
| **PI-CVDs** | **OAPOs** | 10.2 (9.05–11.4) | 11.1 (7.20–17.2) | $I^2$ = 0%, $p$ = 0.69 |
| **Prim hypertension** | **NCVDs** | 2.15 (1.62–2.86) | 2.65 (1.35–5.21) | $I^2$ = 0%, $p$ = 0.58 |
| **Prim anaemia** | **NCVDs** | 1.51 (1.27–1.79) | 1.11 (0.68–1.81) | $I^2$ = 23.4%, $p$ = 0.25 |
| **PI-CVDs** | **NCVDs** | 3.01 (2.10–4.32) | 3.60 (1.59–8.16) | $I^2$ = 0%, $p$ = 0.70 |

**Supplementary Table S9 | STROBE checklist: Checklist of items that should be included in reports of observational studies.**

| | Item No. | Recommendation | Page No.⊥ | Relevant text from manuscript |
|---|---|---|---|---|
| **Title and abstract** | 1 | (*a*) Indicate the study's design with a commonly used term in the title or the abstract | **1** | Maternal cardiovascular and haematological complications alter the risk associations between environmental exposure and adverse pregnancy outcomes |
| | | (*b*) Provide in the abstract an informative and balanced summary of what was done and what was found | **2** | Given China's recent introduction of the "three-child policy" in response to population ageing, safeguarding perinatal health has become an urgent priority. Previous epidemiological research seldom explored the risk factors of maternal cardiovascular and haematological diseases, or its impact on adverse pregnancy outcomes (APO). … |
| **Introduction** | | | | |
| Background/rationale | 2 | Explain the scientific background and rationale for the investigation being reported | **2–3** | Studies have assessed the effects of environmental exposures, including ambient air pollution, heatwave, and green space, on the risk of cardiovascular morbidity and mortality. However, research that focused on the pregnant woman and the neonate, two groups of vulnerable population, is still limited. … |
| Objectives | 3 | State specific objectives, including any prespecified hypotheses | **3** | By systematic analyses of the medical records of pregnant women enrolled in the ZEBRA Chinese maternity cohort, our current study aims to explore: 1) the risk associations between environmental exposures and pregnancy-induced CVDs and APO; 2) the mediating role of maternal cardiovascular symptoms (both primary and pregnancy-induced) in the environment-APO risk association; and 3) the early-stage predictability of APO integrating maternal diagnosis of CVDs. … |
| **Methods** | | | | |
| Study design | 4 | Present key elements of study design early in the paper | **3–4** | The ZEBRA (Zhejiang Environmental and Birth Health Research Alliance) maternity cohort recruited 137,392 Chinese pregnant women nationwide during 2013–2022, among which 121,090 pregnant women were included for epidemiological analyses… |
| Setting | 5 | Describe the setting, locations, and relevant dates, including periods of recruitment, exposure, follow-up, and data collection | **12–13** | From 1 January 2017 to 31 December 2022, ZEBRA maternity cohort enrolled a total of 131,155 parturient women. In addition to the 6,237 participants from the pilot study conducted during 2013–2016… |

| | Item No. | Recommendation | Page No.⊥ | Relevant text from manuscript |
|---|---|---|---|---|
| Participants | 6 | (a) Cohort study—Give the eligibility criteria, and the sources and methods of selection of participants. Describe methods of follow-up<br>Case-control study—Give the eligibility criteria, and the sources and methods of case<br>Cross-sectional study—Give the eligibility criteria, and the sources and methods of selection of participants | **12–14** | ZEBRA maternity cohort collects comprehensive sociodemographic and behavioural features from pregnant women residing in Zhejiang Province, China throughout the study period. … |
| | | (b) Cohort study—For matched studies, give matching criteria and number of exposed and unexposed<br>Case-control study—For matched studies, give matching criteria and the number of controls per case | **N.A.** | |
| Variables | 7 | Clearly define all outcomes, exposures, predictors, potential confounders, and effect modifiers. Give diagnostic criteria, if applicable | **13–14** | In our study, we identified primary (referring to the non-pregnancy-cause diseases throughout all ZEBRA-based studies no matter congenital or acquired) and pregnancy-induced (a type of secondary) cardiovascular complications in all enrolled cohort participants. … |
| Data sources/measurement | 8* | For each variable of interest, give sources of data and details of methods of assessment (measurement). Describe comparability of assessment methods if there is more than one group | **14–15** | In this study, we evaluated maternal exposure to three major environmental factors: ambient $PM_{2.5}$, $O_3$, and green space. … |
| Bias | 9 | Describe any efforts to address potential sources of bias | **15** | The fully adjusted regression model considers other risk factors and potential confounders (such as other environmental exposure factors, socioeconomic characteristics, medical history, and the sex of neonates, … |
| Study size | 10 | Explain how the study size was arrived at | **13** | After excluding participants with missing records for more than 20% of the variables (N=16,091), those with gestational ages less than 20 or greater than 44 weeks (N=39), and pregnant women aged under 18 or over 45 years (N=172), our analyses cover 121,090 pregnant women in total (Fig. 1). There were 124,025 live births, with 116,610 singletons, 3,619 pairs of twins, and 59 sets of triplets, after excluding 802 stillbirths. … |
| Quantitative variables | 11 | Explain how quantitative variables were handled in the analyses. If applicable, | **15** | Maternal exposures were averaged over an 18-month period, from one year before pregnancy until the end of |

| | Item No. | Recommendation | Page No.⊥ | Relevant text from manuscript |
|---|---|---|---|---|
| | | describe which groupings were chosen and why | | the second trimester (the 6th month since conception). |
| Statistical methods | 12 | (*a*) Describe all statistical methods, including those used to control for confounding | **15–16** | Extended Cox proportional hazard regression models with time-varying variables were applied to investigate… |
| | | (*b*) Describe any methods used to examine subgroups and interactions | **15–16** | Additionally, interaction terms between the studied pairs of factors are included in the extended model to assess the effect modification (EM) between risk factors. … |
| | | (*c*) Explain how missing data were addressed | **13** | After excluding participants with missing records for more than 20% of the variables (N=16,091), those with gestational ages less than 20 or greater than 44 weeks (N=39), and pregnant women aged under 18 or over 45 years (N=172), our analyses cover 121,090 pregnant women in total (Fig. 1). There were 124,025 live births, with 116,610 singletons, 3,619 pairs of twins, and 59 sets of triplets, after excluding 802 stillbirths. … |
| | | (d) Cohort study—If applicable, explain how loss to follow-up was addressed<br>Case-control study—If applicable, explain how matching of cases and controls was addressed<br>Cross-sectional study—If applicable, describe analytical methods taking account of sampling strategy | **13** | After excluding participants with missing records for more than 20% of the variables (N=16,091), those with gestational ages less than 20 or greater than 44 weeks (N=39), and pregnant women aged under 18 or over 45 years (N=172), our analyses cover 121,090 pregnant women in total (Fig. 1). There were 124,025 live births, with 116,610 singletons, 3,619 pairs of twins, and 59 sets of triplets, after excluding 802 stillbirths. … |
| | | (*e*) Describe any sensitivity analyses | **19** | We conducted multiple sensitivity analyses to ensure the robustness of our results. These analyses include: … |
| **Results** | | | | |
| Participants | 13* | (a) Report numbers of individuals at each stage of study—eg numbers potentially eligible, examined for eligibility, confirmed eligible, included in the study, completing follow-up, and analysed | **12–13** | From 1 January 2017 to 31 December 2022, ZEBRA maternity cohort enrolled a total of 131,155 parturient women. In addition to the 6,237 participants from the pilot study conducted during 2013–2016… |
| | | (b) Give reasons for non-participation at each stage | **13** | After excluding participants with missing records for more than 20% of the variables (N=16,091), those with gestational ages less than 20 or greater than 44 weeks (N=39), and pregnant women aged under 18 or over 45 years (N=172), our analyses cover 121,090 pregnant women in total (Fig. 1). There were 124,025 live births, with 116,610 singletons, 3,619 pairs of twins, and 59 sets of triplets, after |

| | Item No. | Recommendation | Page No.⊥ | Relevant text from manuscript |
|---|---|---|---|---|
| | | | | excluding 802 stillbirths. … |
| | | (c) Consider use of a flow diagram | **20** | Fig. 1. Recruitment flowchart of ZEBRA maternity cohort participants, 2013–2022. |
| Descriptive data | 14* | (a) Give characteristics of study participants (eg demographic, clinical, social) and information on exposures and potential confounders | **13** | ZEBRA maternity cohort collects comprehensive sociodemographic and behavioural features from pregnant women residing in Zhejiang Province, China throughout the study period. Trained obstetric nurses conduct questionnaire-based face-to-face interviews to record residential address, household registration, ethnicity, education attainment level, smoking habits (active and second-hand), alcohol history, age at current delivery, gravidity, parity, and date of the last menstrual period. Physical examinations are performed to measure the height and weight of the women before conception and at the end of the second trimester. |
| | | (b) Indicate number of participants with missing data for each variable of interest | **13** | After excluding participants with missing records for more than 20% of the variables (N=16,091), those with gestational ages less than 20 or greater than 44 weeks (N=39), and pregnant women aged under 18 or over 45 years (N=172), our analyses cover 121,090 pregnant women in total (Fig. 1). There were 124,025 live births, with 116,610 singletons, 3,619 pairs of twins, and 59 sets of triplets, after excluding 802 stillbirths. |
| | | (c) *Cohort study*—Summarise follow-up time (eg, average and total amount) | **15** | Maternal exposures were averaged over an 18-month period, from one year before pregnancy until the end of the second trimester (the 6th month since conception). |
| Outcome data | 15* | Cohort study—Report numbers of outcome events or summary measures over time<br>Case-control study—Report numbers in each exposure category, or summary measures of exposure<br>Cross-sectional study—Report numbers of outcome events or summary measures | **3–4** | There were 25,544 (21.1%) pregnant women diagnosed with a variety of cardiovascular and haematological complications, including 814 (0.7%) cases of heart disease, 3,760 (3.1%) cases of vascular disease, and 21,935 (18.1%) cases of haematological disease. Pregnancy-induced cardiovascular complications were found in 1,414 (1.2%) cases … |
| Main results | 16 | (*a*) Give unadjusted estimates and, if applicable, confounder-adjusted estimates and their precision (eg, 95% confidence interval). Make clear which confounders were adjusted for and why they were included | **4–10** | Having adjusted for sociodemographic, physiological and behavioural characteristics, medical and disease history, and other environmental exposures (see details in Extended Data Table 3), the risk of pregnancy-induced cardiovascular diseases would increase by 7.3% (hazard ratio, HR=1.073, 95% CI: 1.064–1.082) with each 10-μg/m$^3$ increase in $PM_{2.5}$ exposure, … |

| | Item No. | Recommendation | Page No.⊥ | Relevant text from manuscript |
|---|---|---|---|---|
| | | (b) Report category boundaries when continuous variables were categorised | **S12–14** | Self-directed risk assessment questionnaire for early-stage forecasting of obstetric adverse pregnancy outcomes and neonatal congenital cardiovascular diseases. |
| | | (*c*) If relevant, consider translating estimates of relative risk into absolute risk for a meaningful time period | **3–4** | There were 25,544 (21.1%) pregnant women diagnosed with a variety of cardiovascular and haematological complications, including 814 (0.7%) cases of heart disease, 3,760 (3.1%) cases of vascular disease, and 21,935 (18.1%) cases of haematological disease. Pregnancy-induced cardiovascular complications were found in 1,414 (1.2%) cases … |
| Other analyses | 17 | Report other analyses done—eg analyses of subgroups and interactions, and sensitivity analyses | **17–19** | Ensemble machine learning algorithm for risk prediction… |
| **Discussion** | | | | |
| Key results | 18 | Summarise key results with reference to study objectives | **12–13** | To our knowledge, this is the second Chinese longitudinal maternity cohort study focusing on intrahepatic cholestasis of pregnancy… |
| Limitations | 19 | Discuss limitations of the study, taking into account sources of potential bias or imprecision. Discuss both direction and magnitude of any potential bias | **10–11** | To the best of our knowledge, this is the first Chinese prospective maternity cohort study comprehensively exploring the risk patterns of maternal and neonatal cardiovascular abnormalities during the perinatal period, providing robust primary epidemiological evidence specifically for the East Asian population. … |
| Interpretation | 20 | Give a cautious overall interpretation of results considering objectives, limitations, multiplicity of analyses, results from similar studies, and other relevant evidence | **12** | Last but not the least, our findings may not be representative of the entire Chinese population due to sparse participant enrolment in Northwest China. Therefore, readers should use our results cautiously, particularly when generalising to broader populations, and hence meta-analyses and heterogeneity tests are strongly recommended when more relevant studies come out. It is optimistically hoped that multi-centre collaborative communities will be established soon to enhance the representativeness and generalisability of epidemiological findings. |
| Generalisability | 21 | Discuss the generalisability (external validity) of the study results | **12** | Last but not the least, our findings may not be representative of the entire Chinese population due to sparse participant enrolment in Northwest China. Therefore, readers should use our results cautiously, particularly when generalising to broader populations, and hence meta-analyses and heterogeneity tests are strongly recommended when more relevant studies come out. It is optimistically hoped |

| | Item No. | Recommendation | Page No.⊥ | Relevant text from manuscript |
|---|---|---|---|---|
| | | | | that multi-centre collaborative communities will be established soon to enhance the representativeness and generalisability of epidemiological findings. |
| **Other information** | | | | |
| Funding | 22 | Give the source of funding and the role of the funders for the present study and, if applicable, for the original study on which the present article is based | **36** | This study is funded by the Zhejiang Province Health Innovative Talent Project (A0466), International Cooperation Seed Program of Women's Hospital, Zhejiang University (GH2022B008-01), … |

⊥ Page No. based on the latest version submitted to the journal.
* Give information separately for cases and controls in case-control studies and, if applicable, for exposed and unexposed groups in cohort and cross-sectional studies.
NA indicates items not applicable.
Note: An Explanation and Elaboration article discusses each checklist item and gives methodological background and published examples of transparent reporting. The STROBE checklist is best used in conjunction with this article (freely available on the Websites of PLoS Medicine at http://www.plosmedicine.org/, Annals of Internal Medicine at http://www.annals.org/, and Epidemiology at http://www.epidem.com/). Information on the STROBE Initiative is available at www.strobe-statement.org.

**Supplementary Table S10 | ZEBRA collaborative group full roster.**

*The full roster will be revealed upon acceptance for publication to avoid potential wide conflict of interest.*
